# Cost-Effectiveness Analysis of the mRNA-1345 RSV Vaccine for Older Adults in Italy

**DOI:** 10.64898/2026.08.17.26360570

**Authors:** Mariia Dronova, Camille Moyon, Lukasz Pyrek, Katherine Hicks, Ziyi Xiao, Filippo Rumi, Chiara de Waure, Stefan Scholz, Parinaz Ghaswalla

**Affiliations:** Inizio Ignite Putnam, Paris, France; Inizio Ignite Putnam, Krakow, Poland; RTI Health Solutions, Durham, NC, USA; ALTEMS Advisory, Spin-off of Università Cattolica del Sacro Cuore, Rome, Italy; Department of Medicine and Surgery, University of Perugia; Moderna Germany GmbH, Munich, Germany; Moderna Inc, Cambridge, MA, USA

**Keywords:** Cost-effectiveness, Clinical value, Economic value, mRNA, Respiratory syncytial virus, Vaccines

## Abstract

**Introduction:** Respiratory syncytial virus (RSV) is an important cause of respiratory disease in older adults and adults with chronic medical conditions, contributing substantially to the healthcare burden in Italy. The availability of effective RSV vaccines provides an opportunity to reduce RSV-related morbidity, mortality, and healthcare costs in populations at high risk of severe disease. This study evaluates the potential public health impact and cost-effectiveness of vaccination using mRNA-1345 administered as a single dose compared with no vaccination in Italian high-risk adults aged 60-74 years and all adults aged ≥75 years.

**Methods:** A static decision-analytic model was developed to project clinical and economic outcomes over a 5-year time horizon. Economic outcomes were evaluated from the Italian National Health Service (Servizio Sanitario Nazionale, SSN) perspective. Model inputs were informed by the most recent Italian epidemiological, clinical, and economic evidence, supplemented by published international data when necessary. Deterministic, probabilistic, and scenario analyses were conducted to assess the impact of uncertainty in model inputs and assumptions on the study results.

**Results:** Vaccination with mRNA-1345 in high-risk adults aged 60-74 years and all adults aged ≥75 years was projected to avert over 19,800 hospitalizations, 4,000 emergency department visits, 381,000 outpatient visits, 6,000 RSV-attributable deaths, and 212,000 antibiotic prescriptions compared with no vaccination over a 5-year period. The total incremental cost of €1,143 million and the additional 47,477 QALYs gained resulted in an ICER of €24,078, which was below the commonly referenced willingness to-pay range of €33,000- 40,000 per QALY gained. Sensitivity analyses confirmed robustness of the analysis results.

**Conclusions:** Vaccination with mRNA-1345 is a cost-effective strategy for the prevention of RSV in high-risk adults aged 60-74 years and all adults ≥75 years in Italy and has the potential to provide substantial public health benefits.

**Key Summary Points:** *Why carry out this study?:* The clinical and economic burden of respiratory syncytial virus (RSV) in high-risk and older adults is significant. RSV vaccines have received regulatory authorization in Europe, and national recommendations or immunization programs for older adults have been introduced in several countries including Germany, France, Spain and the United Kingdom. In Italy, RSV vaccination for older adults has not yet been implemented as a uniform national program, however such a program has been endorsed by several scientific societies. The study objective was to estimate the potential public health impact and cost-effectiveness of vaccination using mRNA-1345 administered as a single dose compared with no vaccination in Italian high-risk adults aged 60-74 years and all adults aged ≥75 years, in the 5-year period following vaccination.

*What was learned from the study?:* Vaccination of high-risk adults aged 60-74 years and all adults aged ≥75 years with mRNA- 1345 was projected to prevent more than 19,800 hospitalizations, 4,000 ED visits, 381,000 outpatient visits, 6,000 RSV-attributable deaths, and 212,000 antibiotic prescriptions (up to 21% reduction across outcomes), in comparison to no vaccination, over a 5-year period. This analysis suggests the mRNA-1345 vaccine is a cost-effective strategy for the prevention of RSV in high-risk adults in Italy and has the potential for substantial public health benefit. Our findings provide evidence to inform national immunization policy and reimbursement decisions, and support the implementation of an RSV vaccination program for high-risk adults aged 60-74 years and all adults aged ≥75 years in Italy.

**Plain Language Summary:** Respiratory syncytial virus (RSV) is a common virus that causes lung infections. It can be especially dangerous for older adults and people with health conditions like asthma, heart disease, or diabetes. Each year, RSV causes many hospitalizations and deaths in Italy. A new vaccine called mRNA-1345 was recently approved to protect individuals aged 18 years and older against severe RSV disease. This study looked at whether vaccinating the most vulnerable adults, particularly people aged 60-74 years with chronic health conditions and everyone aged 75 years and older, would help reduce the burden of RSV and provide good value for money for the healthcare system. Researchers used a mathematical model to compare what might happen over 5 years with and without vaccination. They found that using a single-dose vaccination strategy could prevent over 19,800 hospitalizations and more than 6,000 deaths over the following 5 years. Although vaccination requires investments, the health benefits are estimated to be large enough for mRNA-1345 to provide good value for money for the Italian healthcare system.

## Introduction

Respiratory syncytial virus (RSV) is a highly contagious common virus which can cause upper and lower respiratory tract infections, which if severe can lead to complications including exacerbation of pre-existing conditions, hospitalization and death (1, 2). Populations characterized as immunocompromised, elderly, or those with chronic comorbidities (e.g., asthma, diabetes mellitus, cardiovascular diseases) are at an increased risk of severe disease. The clinical and economic burden of RSV is substantial, affecting 64 million people and causing 160,000 deaths worldwide annually (3). RSV burden is unequally distributed across age groups, with children and older adults experiencing a disproportionate share of RSV-associated hospitalizations. With effective preventive strategies now available for infants, increasing attention has shifted towards addressing the burden of RSV in older adults. The RSV Consortium in Europe estimated that adults ≥65 years account for 92% of RSV-associated hospitalizations among all adult patients (4). In Italy, a considerable burden is also evident in absolute terms: among adults ≥60 years, Savic et al. estimated approximately 290,000 RSV cases, 26,000 hospitalizations, and 1,800 RSV-attributable in-hospital deaths annually (5). This clinical burden translates into a substantial economic burden. The mean direct healthcare cost per patient ≥60 years during the index hospitalization and 12-month follow-up has been estimated as €11,599, with hospitalization, prescription, and outpatient services accounting for 79%, 16%, and 5% of this cost, respectively (6).

Furthermore, the overall clinical and economic burden of RSV in Italy is likely underestimated, partially due to limitations in both clinical case ascertainment and diagnostic testing. Much of the available Italian evidence has historically been generated within influenza surveillance frameworks using influenza-like-illness or fever-based criteria (2, 7). While these systems provide valuable information on seasonal respiratory virus circulation, such approaches may under-ascertain RSV burden, particularly among patients with afebrile presentation, which is relatively common in adults (8). In addition, virological confirmation has commonly relied on testing of upper-respiratory tract swabs (7), which has suboptimal sensitivity for RSV detection in adults (9).

Consistent with these limitations, under-ascertainment of RSV burden in older adults has been widely recognized, with reported hospitalization estimates likely representing only a fraction of the true burden (2, 9–11). Although evidence remains limited in Italy, a recent modeling study by Cozzolino et al. (12) suggests that the true RSV-associated hospitalization burden in older adults may be up to 8-fold higher than routinely reported, depending on age.

In the absence of a specific curative treatment for RSV, clinical management of severe RSV disease remains primarily supportive, which underscores the global need for preventive strategies, such as vaccination, to reduce RSV-associated morbidity and healthcare burden (13). Since 2023, three vaccines have been approved for use by the European Medicines Agency to protect adults aged ≥18 years against RSV-related lower respiratory tract disease (RSV-LRTD), including RSVPreF (Abrysvo), RSVPreF3 (Arexvy) and mRNA-1345 (mRESVIA) (14–16). However, although RSV vaccines for adults are available in Italy, a national RSV vaccination program for older adults has not yet been implemented. Several Italian scientific societies have endorsed such a program, with the Italian Society of Hygiene, Preventive Medicine and Public Health and the Italian Society of Infectious and Tropical Diseases jointly recommending vaccination for adults aged ≥60 years with comorbidities and all adults aged ≥75 years (17, 18). In contrast, several European countries, including Germany, France, Spain, and the United Kingdom, have already adopted national RSV vaccination programs or recommendations for older adults (1, 19). As Italy considers the implementation of a national RSV vaccination program for older adults, robust evidence on the clinical and economic value of the available vaccines is needed to support evidence-based decision-making and healthcare resource allocation. Recent Italian economic evaluations have demonstrated the cost-effectiveness of RSVPreF3 and bivalent prefusion F vaccination in older adults (20–23). However, to our knowledge, no economic evaluation of mRNA-1345 has been published for the Italian setting to date.

In this study, we developed a decision-analytic model to estimate the potential public health impact and cost-effectiveness of vaccination with mRNA-1345 administered as a single dose compared with no vaccination in Italian high-risk adults aged 60-74 years and all adults aged ≥75 years (hereafter, “the target population”). Economic outcomes were evaluated from the Italian National Health Service (Servizio Sanitario Nazionale, SSN) perspective.

## Methods

### Model Overview

A static decision-analytic model was developed to estimate the public health impact and cost-effectiveness of one-time vaccination with mRNA-1345 compared to a strategy of no vaccination. The model estimated clinical outcomes including cases of RSV-related acute respiratory disease (RSV-ARD), RSV-related lower respiratory tract disease (RSV-LRTD), level of medical care, deaths attributable to RSV, antibiotic use and quality-adjusted life years (QALYs). Cost-effectiveness was assessed using the incremental cost-effectiveness ratio (ICER) calculated as the incremental cost per QALY gained. As no explicit willingness-to-pay (WTP) threshold has been established in Italy, ICERs were evaluated against two reference WTP values: €33,000 and €40,000 per QALY gained (24). The reference value of €33,000 per QALY gained was informed by the mean ICER of medicines reimbursed by the SSN, based on negotiated net prices in Agenzia Italiana del Farmaco (AIFA) reimbursement decisions between 2016 and 2021 (mean ICER was estimated at €33,004 per QALY). A value of €40,000 per QALY gained was considered as an alternative, higher reference value (24).

Figure 1 presents the decision tree applied annually in the model. Table 1 presents key model inputs and sources. A comprehensive list of model inputs is provided in Supplementary materials, Table S4.

**Figure 1.**
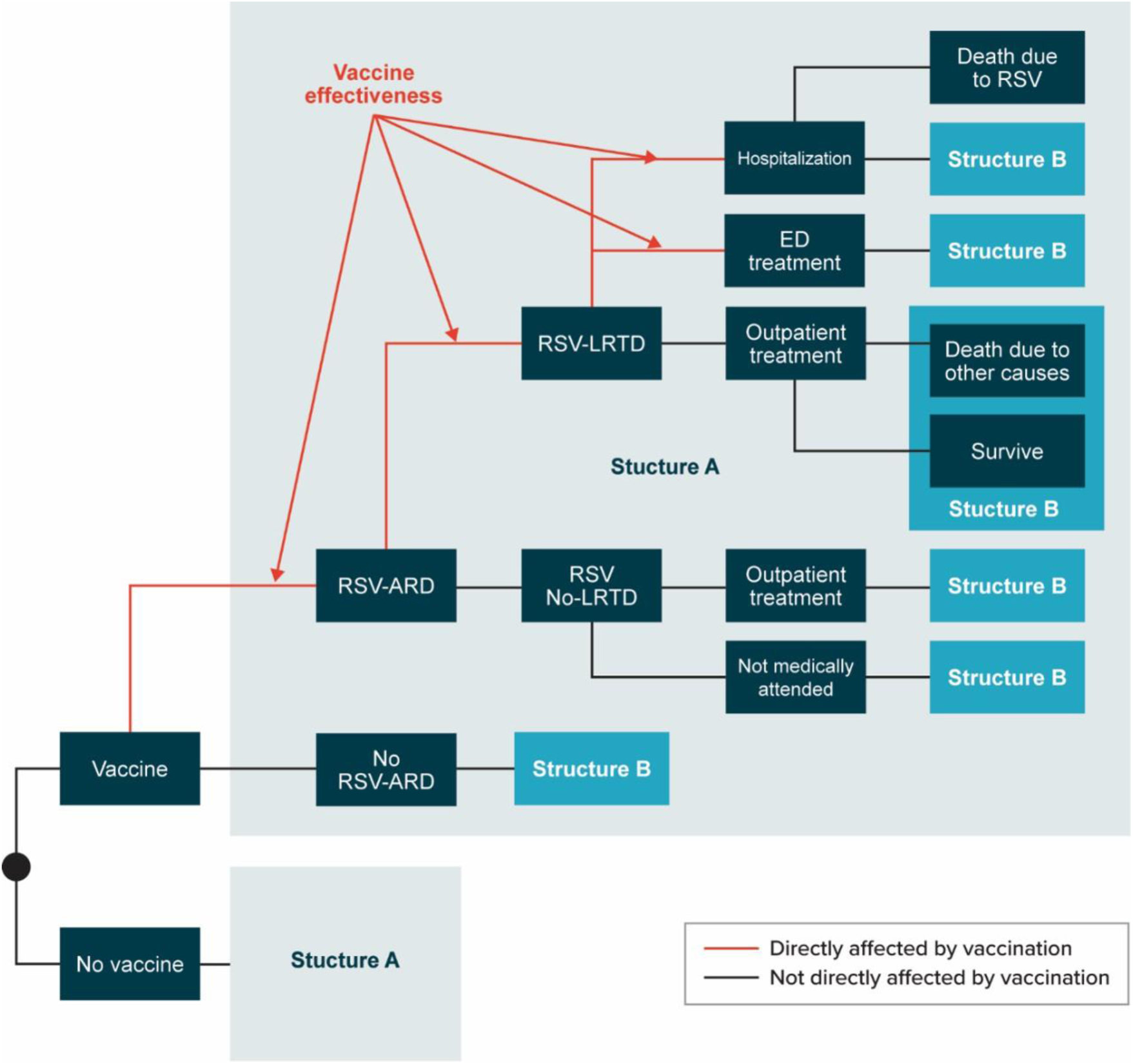
Model Structure. ED, emergency department, RSV, respiratory syncytial virus, RSV-ARD, RSV-related acute respiratory disease, RSV-LRTD, RSV-related lower respiratory tract disease.

**Table 1.** Key model inputs ED, emergency department, HRQoL, health-related quality of life, QALY, quality-adjusted life-year, RSV, respiratory syncytial virus, RSV-ARD, RSV-related acute respiratory disease, RSV-LRTD, RSV-related lower respiratory tract disease, VE, vaccine efficacy.

| Model input parameter | Base-case value | Data source |
| --- | --- | --- |
| Settings |  |  |
| Discount rates for costs and outcomes | 3.0% | AIFA HTA Guidelines (55) |
| Population |  |  |
| Italian population size, by age group |  |  |
| 60-64 years | 4,566,052 | Italian National Institute of Statistics (ISTAT) (25) |
| 65-69 years | 3,831,068 |  |
| 70-74 years | 3,278,863 |  |
| 75-79 years | 3,095,916 |  |
| 80-84 years | 2,103,261 |  |
| 85+ years | 2,511,392 |  |
| Percentage of individuals at high risk, by age group* |  |  |
| 60-64 years | 48.9% | PASSI d'Argento surveillance system (26), assuming the same percentage for individuals aged 60-64 years as reported for those aged 65-74 years |
| 65-69 years | 48.9% |  |
| 70-74 years | 48.9% |  |
| 75-79 years | 62.5% |  |
| 80-84 years | 62.5% |  |
| 85+ years | 71.6% |  |
| Vaccination coverage rate and effectiveness |  |  |
| mRNA-1345 vaccine coverage, all age and risk groups | 50.0% | Assumption based on coverage reported for influenza vaccination in 2024-2025 season (40) |
| Estimated average annual vaccine efficacy against RSV-ARD |  |  |
| Initial VE, 1 month post-vaccination | 69.2% | Evidence from clinical trial data (44), projection over 5 years |
| Year 1 post-vaccination | 53.9% |  |
| Year 2 post-vaccination | 39.2% |  |
| Year 3 post-vaccination | 31.5% |  |
| Year 4 post-vaccination | 25.9% |  |
| Year 5 post-vaccination | 21.4% |  |
| Estimated average annual vaccine efficacy against RSV-LRTD (hospitalized and non-hospitalized cases) |  |  |
| Initial VE, 1 month post-vaccination | 76.9% | Evidence from clinical trial data (44), projection over 5 years |
| Year 1 post-vaccination | 60.7% |  |
| Year 2 post-vaccination | 43.9% |  |
| Year 3 post-vaccination | 34.5% |  |
| Year 4 post-vaccination | 27.5% |  |
| Year 5 post-vaccination | 21.7% |  |
| Vaccine-related adverse events, all age and risk groups |  |  |
| Adverse event rates, per 100,000 doses |  |  |
| Grade 3 local | 1,426 | Wilson et al. (27) |
| Grade 3 systemic | 1,133 |  |
| Adverse event costs |  |  |
| Grade 3 local | €28.56 | AIFA transparency lists (63), Capri et al. (47), ISTAT (57, 58) |
| Grade 3 systemic | €28.53 | AIFA transparency lists (63), Ministry of Health decree of November 25, 2024 (64), ISTAT (58) |
| QALY loss per adverse event |  |  |
| Grade 3 local | 0.0003 | Estimated based on Prosser et al. (53) |
| Grade 3 systemic | 0.0011 |  |
| RSV epidemiology and clinical outcomes (without vaccination) |  |  |
| Percentage experiencing RSV-ARD, all age and risk groups | 5.7% | Korsten et al. (31) |
| Percentage experiencing RSV-LRTD, by age group |  |  |
| 60-64 years | 8.8% | Estimated based on: Korsten et al. (31), Fleming et al. (33), McLaughlin et al. (11) and Osei-Yeboah et al. (4) |
| 65-69 years | 9.6% |  |
| 70-74 years | 9.6% |  |
| 75-79 years | 20.9% |  |
| 80-84 years | 20.9% |  |
| 85+ years | 21.8% |  |
| Percentage of RSV-LRTD patients requiring hospitalization, non-high-risk individuals, by age group |  |  |
| 60-64 years | 3.1% | Estimated based on: Fleming et al. (33), McLaughlin et al. (11), Osei-Yeboah et al. (4, 34), and Italian PASSI d'Argento surveillance system (26) |
| 65-69 years | 5.3% |  |
| 70-74 years | 5.3% |  |
| 75-79 years | 18.7% |  |
| 80-84 years | 18.7% |  |
| 85+ years | 18.7% |  |
| Percentage of RSV-LRTD patients requiring hospitalization, high-risk individuals, by age group |  |  |
| 60-64 years | 8.8% | Estimated based on: Fleming et al. (33), McLaughlin et al. (11), and Osei-Yeboah et al. (34) |
| 65-69 years | 15.2% |  |
| 70-74 years | 15.2% |  |
| 75-79 years | 18.7% |  |
| 80-84 years | 18.7% |  |
| 85+ years | 18.7% |  |
| Percentage of RSV-LRTD patients requiring ED treatment, all age and risk groups | 3.5% | Baglivo et al. (35) |
| Percentage of RSV-No LRTD patients requiring outpatient treatment, all age and risk groups | 55.7% | Dal Negro et al. (36) |
| Mortality due to RSV-LRTD requiring hospitalization, including in-hospital and 1-month post-discharge mortality, by age group |  |  |
| 60-64 years | 8.0% | Estimated using the data reported by Puggina et al. (2) and adjusted to account for all-cause mortality (37) |
| 65-69 years | 17.8% |  |
| 70-74 years | 17.8% |  |
| 75-79 years | 34.6% |  |
| 80-84 years | 34.6% |  |
| 85+ years | 34.6% |  |
| Mortality due to RSV-LRTD requiring ED treatment, outpatient treatment, or no treatment, all age and risk groups | 0.0% | Assumption |
| Percentage of RSV-ARD cases with antibiotics use, by level of medical care, all age and risk groups |  |  |
| Hospitalization | 61.2% | Puggina et al. (2) |
| ED treatment | 77.0% | Santus et al. (39) |
| Outpatient treatment | 51.6% | Bracaloni et al. (38) |
| No treatment | 0.0% | Assumption |
| **Direct costs, all age and risk groups** |  |  |
| Acquisition cost of mRNA-1345 | €180 | Assumption in line with vaccine list price assumptions used in previously published Italian economic evaluations (21, 22) |
| Administration cost of mRNA-1345 | €7.47 | Capri et al. (47), ISTAT (58) |
| Cost per case of RSV requiring hospitalization | €4,282 | Estimated based on mean cost of DRGs 079, 080, 089, 090, 092, 093, 096, and 097 (46), ISTAT (57, 58) |
| Cost per case of RSV requiring ED visit | €285 | Calabrò et al. (20), ISTAT (58) |
| Cost per case of RSV requiring outpatient visit | €98 | Calabrò et al. (20), ISTAT (58) |
| **HRQoL** |  |  |
| Baseline utility, by age group |  |  |
| 60-64 years | 0.907 | Scalone et al. (59) |
| 65-69 years | 0.892 |  |
| 70-74 years | 0.892 |  |
| 75-79 years | 0.853 |  |
| 80-84 years | 0.853 |  |
| 85+ years | 0.853 |  |
| QALY loss per level of medical care, all age and risk groups |  |  |
| Hospitalization | 0.0193 | Hutton et al. (60, 61) |
| ED treatment | 0.0185 | Assumption |
| Outpatient treatment | 0.0185 | Hutton et al. (60, 61) |
| No treatment | 0.0093 | Assumption based on Moghadas et al. (62) |

\* Individuals with at least one chronic condition were defined according to the PASSI d'Argento surveillance system as those with cardiovascular disease, stroke or cerebral ischemia, cancer (including leukemia and lymphoma), chronic respiratory diseases, diabetes, chronic liver disease/cirrhosis, or renal failure.

### Population

The population for the base-case analysis included adults aged 60-74 years at high-risk due to chronic medical conditions and all adults aged ≥75 years, in line with recent recommendations from Italian scientific societies (1, 19). The following conditions were considered as risk factors for severe RSV: cardiovascular, lung, neurological, hematological, liver diseases, chronic kidney disease, diabetes mellitus, severe obesity, cancer, and decreased immune function (19).

Age-specific estimates of the general population size were informed by the Italian National Institute of Statistics (ISTAT) (25), the official source of national demographic statistics in Italy. In addition, the proportions of individuals classified as non-high-risk and high-risk were derived from the Italian PASSI d’Argento surveillance system, based on nationally representative data on the proportion of adults with one chronic condition (26).

### Intervention

In this study, vaccination with mRNA-1345, administered as a single dose, is compared with no vaccination for the target population. The mRNA-1345 vaccine was assumed effective against RSV-ARD and RSV-LRTD, based on the clinical trial data (27). The vaccine effectiveness for hospitalization and emergency department (ED) treatment was assumed to be the same as the clinical trial efficacy against RSV-LRTD. This assumption is supported by real world evidence demonstrating that vaccine effectiveness against medically attended RSV disease, including hospitalization, ED visits, and outpatient visits, is broadly consistent with the efficacy observed against RSV-LRTD in clinical trials (27–30). Therefore, within the model structure, preventing a case of RSV-ARD or RSV-LRTD was assumed to prevent all RSV-related downstream health consequences.

### RSV Epidemiology and Clinical Outcomes

The annual incidence of RSV-ARD infection without vaccination was derived from Korsten et al., who conducted a prospective cohort study of community-dwelling older adults across Europe to estimate the incidence and burden of RSV infection (31). The same RSV incidence pattern was applied for each year of the time horizon, recognizing that historically, the seasonal patterns have varied across years. The proportion of RSV-ARD infections occurring each month in Italy was based on 2023/2024 report of Istituto Superiore di Sanità (32).

The proportion of patients with RSV-ARD who develop LRTD is uncertain and rarely reported, likely due to variability in LRTD case definitions, changes in the standard of care over time, and reliance on hospitalization as a main measure of RSV burden. Data for the high-risk population are even more limited. Therefore, it was considered appropriate to calibrate the proportion of RSV-LRTD, considering the available data on hospitalization rates. Data derivation and calibration was conducted as a stepwise procedure, described in detail in Supplementary materials. Briefly, a proportion of patients with RSV-ARD who develop LRTD was calibrated to reproduce the age-specific hospitalization rate estimates reported by Osei-Yeboah et al. for the general population in Italy (4). This calibration also incorporated published estimates of the probability of hospitalization among patients with RSV-LRTD (33), adjusted for under-ascertainment using a correction factor of 1.5 (11). Given the limited evidence available for high-risk adults, the increased risk associated with underlying medical conditions was applied only to the probability of hospitalization following RSV-LRTD, applying a conservative relative risk (RR) of 1.5 informed by the lowest estimate reported by Osei-Yeboah et al. (34).

The percentage of RSV-LRTD cases that require ED visits and RSV-No LRTD cases that require outpatient treatment were derived using data reported by Baglivo et al. and Dal Negro et al., respectively (35, 36), with the same values applied across age and risk groups.

It was assumed that RSV-related mortality occurred only among hospitalized patients. The percentage of hospitalized RSV-LRTD cases resulting in death was estimated using the combined in-hospital and 1-month post-discharge mortality data reported by Puggina et al., which was adjusted to account for all-cause mortality, to avoid double-counting (2, 37). Further details on the data derivation are provided in the Supplementary Materials.

The percentage of RSV-ARD cases receiving antibiotics by level of medical care was informed by the published Italian studies (2, 38, 39).

### Vaccination Coverage Rate and Efficacy

RSV vaccination coverage rate was assumed to be 50% across all age and risk groups, which was in line with the coverage reported for influenza vaccination (52.5% for general population aged ≥65 years, 2024-2025 season) (40).

Vaccine efficacy against RSV-ARD and RSV-LRTD over time was modeled using nonlinear waning curves. A nonlinear waning profile was assumed to reflect the efficacy and immunogenicity evidence from clinical trial data, including sustained vaccine efficacy through 24 months post-vaccination, together with evidence that RSV neutralizing antibody titers remained robust and well above baseline levels between 12 and 24 months post-vaccination, supported by durable cell-mediated immunity across age and risk groups and by emerging evidence linking antibody levels with protection (41–43).

To estimate the duration of protection following mRNA-1345 vaccination, nonlinear models were fitted to monthly vaccine efficacy estimates from the extended clinical efficacy analysis, which demonstrated sustained protection through the 24-month follow-up period (44). The model used the logarithm of time as the independent variable and monthly log incidence rate ratio as the dependent variable. Further details on the applied methodology are provided in Supplementary Materials.

Modeled vaccine efficacy against RSV-ARD and RSV-LRTD was projected up to 60 months, with the rate of waning slowing beyond 24 months. Consistent with recommendations from the ISPOR-SMDM Modeling Good Research Practices Task Force, the base-case analyses used a 5-year time horizon to capture the full benefits of vaccination (45).

Annual average vaccine efficacy derived from the fitted waning functions and used in the cost-effectiveness model are presented in Table 1. Further, monthly vaccine efficacy measures were weighted by incidence seasonality patterns and assuming that RSV vaccination would be given in October, prior to the typical start of the RSV season in Italy.

### Costs

Economic outcomes were evaluated from the perspective of the Italian National Health Service (Servizio Sanitario Nazionale, SSN); therefore, only direct healthcare costs were included in the base-case analysis. The direct cost of RSV-related hospitalization applied in the model was estimated using the mean cost of Diagnosis-Related Groups (DRGs) 079, 080, 089, 090, 092, 093, 096, and 097, based on the most recent Italian DRG tariffs published by the Ministry of Health (46). The costs of RSV cases requiring ED visits and outpatient management were sourced from Calabrò et al., who evaluated vaccination with the adjuvanted RSVPreF3 vaccine in Italian adults using country-specific healthcare costs (20). A cost of €0 was assumed for patients who did not require any medical treatment. The unit cost of the mRNA-1345 vaccine was assumed to be €180, consistent with vaccine list price assumptions used in previously published Italian economic evaluations (21, 22). The administration cost was sourced from Capri et al., who conducted a cost-effectiveness analysis comparing seasonal influenza vaccination strategies in Italian adults aged ≥65 years from the SSN perspective (47).

Indirect costs were excluded from the base-case analysis but were considered in a dedicated scenario analysis. Indirect costs included the loss of productive time from missed work due to receiving the mRNA-1345 vaccine or for RSV cases. The cost of lost productivity was calculated using data from the ILOSTAT database (International Labour Organization), ISTAT database and published literature by multiplying the percentage of adults in the labor force by mean hourly income for the total population and the average number of hours worked per day, then further multiplying by the days expected to be lost from work due to either receiving the vaccination or RSV infection (2, 48–53). Productivity losses due to absenteeism for RSV cases by level of medical care were also included. Productivity losses due to premature mortality were estimated using the human capital approach (37, 50–52, 54–56).

All costs were inflated to 2025 euros using “all items” component of the Italian Consumer Price Index (CPI) for the whole nation published by ISTAT (57, 58).

### QALYs

QALY losses were incurred as a result of RSV morbidity and mortality and vaccine-related adverse events. Age-specific population utility values were used to calculate the QALYs lost due to death from RSV, discounted to present value. Baseline utility values were calculated as weighted averages of age- and sex-specific mean EuroQol 5-Dimension 5- Level (EQ-5D-5L) utility values reported by Scalone et al. (59). The values provided by Hutton et al. were used as the base-case estimates for the number of QALYs lost by patients requiring hospitalization, ED, and outpatient treatment, assuming that those seeking ED treatment would experience QALY losses equivalent to those seeking outpatient treatment (60, 61). Consistent with the study by Moghadas et al. (62), it was assumed that patients who did not seek medical care would experience 50% of the QALY losses of patients receiving outpatient treatment. QALY losses for vaccine adverse events were estimated using the assumptions described by Prosser et al. (53).

### Outcomes

Clinical outcomes captured to estimate public health burden include cases of RSV-LRTD and RSV-No LRTD disaggregated by level of medical care required (e.g., hospitalization; ED treatment; outpatient treatment; no treatment), deaths attributable to RSV, QALYs lost, and antibiotic use. Economic outcomes captured for estimating cost-effectiveness included RSV-related direct medical costs, vaccine related costs (including unit cost, administration, and adverse events). Indirect costs were considered in a dedicated scenario analysis only (including productivity loss due to vaccination and adverse events, and productivity loss due to RSV-related morbidity and mortality).

### Scenario and Sensitivity Analysis

Eleven additional scenario analyses were conducted to assess the impact of vaccination under different assumptions for the target subgroup, vaccination coverage, RSV case severity, associated cost and QALY loss, a shorter time horizon, and analysis perspective, as summarized in Table 2.

**Table 2.**
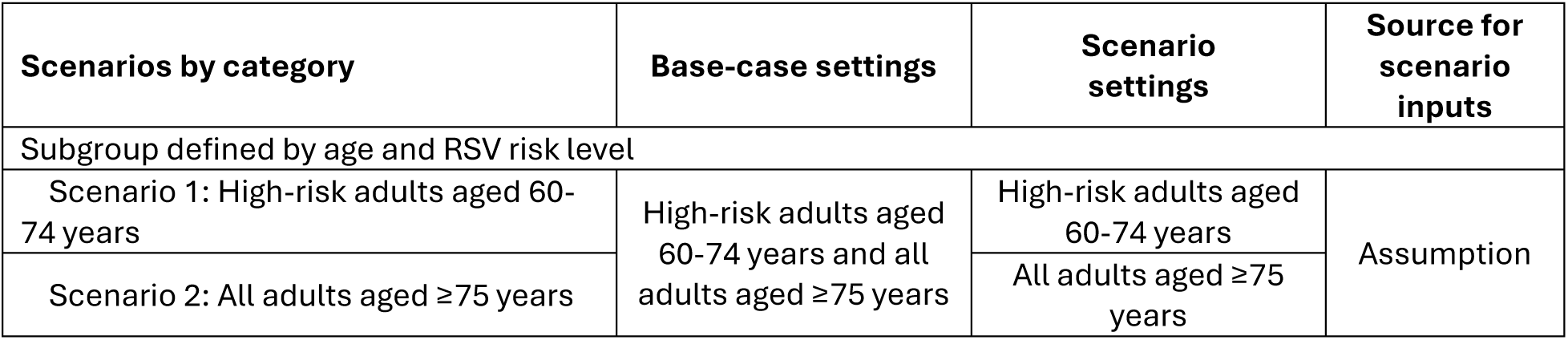
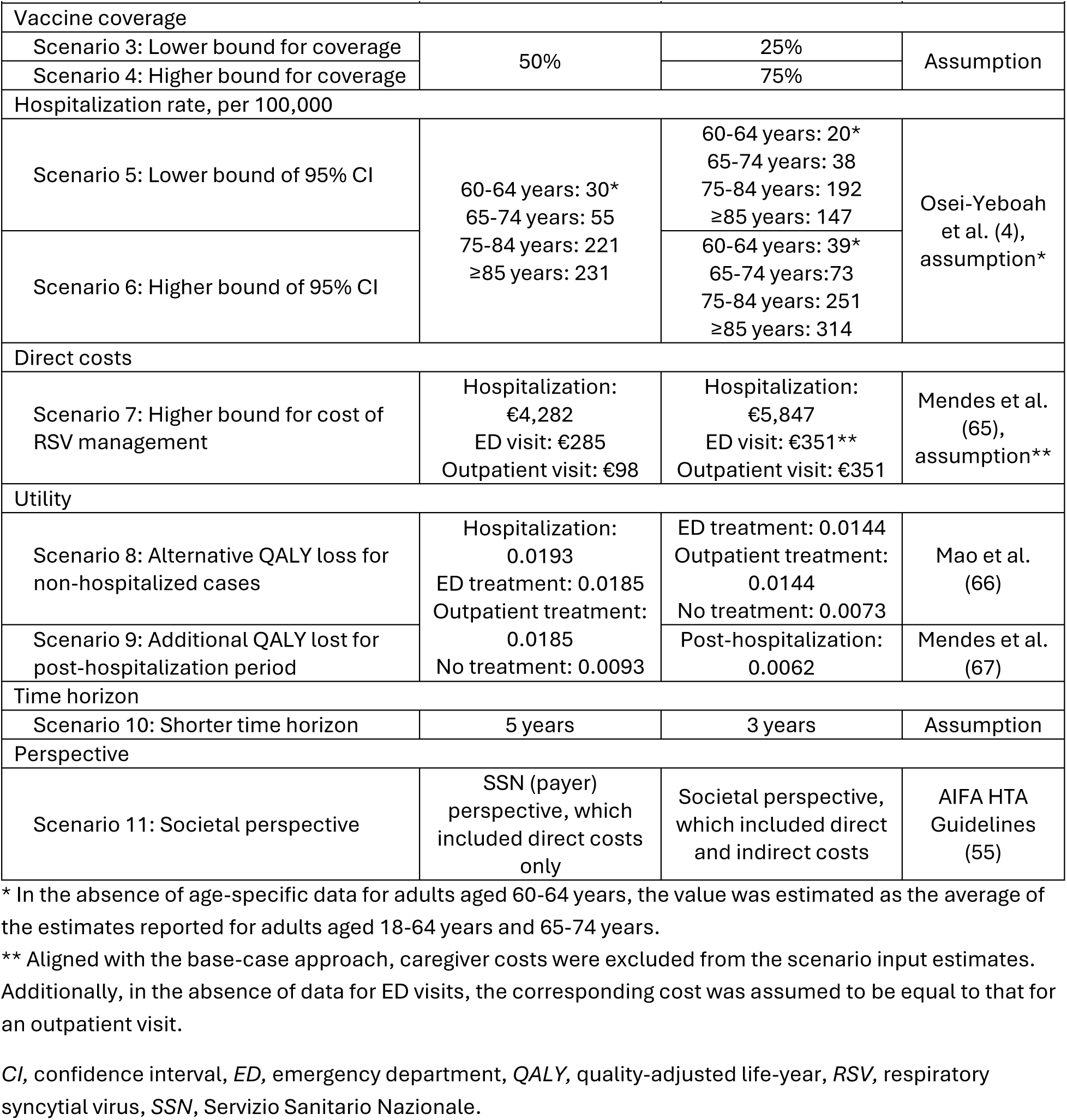
List of scenario analyses CI, confidence interval, ED, emergency department, QALY, quality-adjusted life-year, RSV, respiratory syncytial virus, SSN, Servizio Sanitario Nazionale.

| Scenarios by category | Base-case settings | Scenario settings | Source for scenario inputs |
| --- | --- | --- | --- |
| Subgroup defined by age and RSV risk level |  |  |  |
| Scenario 1: High-risk adults aged 60-74 years | High-risk adults aged 60-74 years and all adults aged ≥75 years | High-risk adults aged 60-74 years | Assumption |
| Scenario 2: All adults aged ≥75 years |  | All adults aged ≥75 years |  |
| Vaccine coverage |  |  |  |
| Scenario 3: Lower bound for coverage | 50% | 25% | Assumption |
| Scenario 4: Higher bound for coverage |  | 75% |  |
| Hospitalization rate, per 100,000 |  |  |  |
| Scenario 5: Lower bound of 95% CI | 60-64 years: 30*<br>65-74 years: 55<br>75-84 years: 221<br>≥85 years: 231 | 60-64 years: 20*<br>65-74 years: 38<br>75-84 years: 192<br>≥85 years: 147 | Osei-Yeboah et al. (4), assumption* |
| Scenario 6: Higher bound of 95% CI |  | 60-64 years: 39*<br>65-74 years: 73<br>75-84 years: 251<br>≥85 years: 314 |  |
| Direct costs |  |  |  |
| Scenario 7: Higher bound for cost of RSV management | Hospitalization: €4,282<br>ED visit: €285<br>Outpatient visit: €98 | Hospitalization: €5,847<br>ED visit: €351**<br>Outpatient visit: €351 | Mendes et al. (65), assumption** |
| Utility |  |  |  |
| Scenario 8: Alternative QALY loss for non-hospitalized cases | Hospitalization: 0.0193<br>ED treatment: 0.0185<br>Outpatient treatment: 0.0185<br>No treatment: 0.0093 | ED treatment: 0.0144<br>Outpatient treatment: 0.0144<br>No treatment: 0.0073 | Mao et al. (66) |
| Scenario 9: Additional QALY lost for post-hospitalization period |  | Post-hospitalization: 0.0062 | Mendes et al. (67) |
| Time horizon |  |  |  |
| Scenario 10: Shorter time horizon | 5 years | 3 years | Assumption |
| Perspective |  |  |  |
| Scenario 11: Societal perspective | SSN (payer) perspective, which included direct costs only | Societal perspective, which included direct and indirect costs | AIFA HTA Guidelines (55) |

\* In the absence of age-specific data for adults aged 60-64 years, the value was estimated as the average of the estimates reported for adults aged 18-64 years and 65-74 years.
\*\* Aligned with the base-case approach, caregiver costs were excluded from the scenario input estimates. Additionally, in the absence of data for ED visits, the corresponding cost was assumed to be equal to that for an outpatient visit.

All scenario and sensitivity analyses, with the exception of a Scenario 11, adopted the perspective of the Italian National Health Service (SSN), and considered only direct healthcare costs. Scenario 11 adopted a societal perspective and included both direct and indirect costs.

For the deterministic sensitivity analysis (DSA), parameters were varied in groups to determine the key drivers of the model. In each group of parameters, the individual inputs were varied in line with the ranges reported in the base-case data sources or varied by +/- 20% if the literature data were not available. Groups of parameters stratified by age such as, e.g., the percentage experiencing RSV-ARD or percentage of RSV cases experiencing LRTD were varied simultaneously in the DSA to improve interpretability. The groups of parameters and the values used in the DSA are shown in Table S4. The impact of applying the upper and lower bounds for each group was presented on a tornado graph for the most influential parameter groups (sorted by impact, with the most impactful groups at the top) and assessed considering the commonly referenced WTP range of €33,000-€40,000 per QALY gained.

Probabilistic sensitivity analyses (PSA) were conducted to account for the joint uncertainty of the underlying parameter estimates. Monte Carlo simulation (random sampling of parameters values from pre-defined distributions) was used. Distributions were chosen in line with the recommendations given by Briggs et al. (68). The distribution parameters were informed by the CIs reported in the literature or calibrated to reflect the +/-20% variation assumed if the literature data were not available. A total of 2,000 iterations were run. The parameters that were varied probabilistically are summarized in Table S4. The outcomes of this analysis included the mean incremental costs, mean incremental QALYs, and the probabilistic ICER calculated as the ratio of the mean incremental costs to the mean incremental QALYs across all Monte Carlo simulations. The results were also presented on a cost-effectiveness plane with two reference WTP values (€33,000 and €40,000 per QALY gained), and a cost-effectiveness acceptability curve (CEAC) displaying the probability of being cost-effective across a broad range of WTP values.

### Ethical Approval

Data for this study were collected and analyzed from published research and did not require additional ethical review. No new studies with human participants or animals were performed by any of the authors.

## Results

### Base-Case Analysis

The population for the base-case analysis included adults aged 60-74 years at high-risk due to chronic medical conditions and all adults aged ≥75 years. Assuming 50% vaccination coverage, approximately 7 million individuals out of a total eligible population of approximately 13 million would be vaccinated with mRNA-1345. Without vaccination, the model predicted more than 96,000 hospitalizations, 19,000 ED visits, 2,000,000 outpatient visits, 30,000 deaths attributable to RSV, and 1,000,000 antibiotic prescriptions, over a 5-year time horizon. Vaccination with a single dose of mRNA-1345 was projected to prevent more than 19,800 hospitalizations, 4,000 ED visits, 381,000 outpatient visits, 6,000 deaths, and 212,000 antibiotic prescriptions (18% to 21% reduction across outcomes), in comparison to no vaccination (Table 3). Overall, the projected health benefits of vaccination yielded 47,477 QALYs gained over the 5-year time horizon.

**Table 3.**
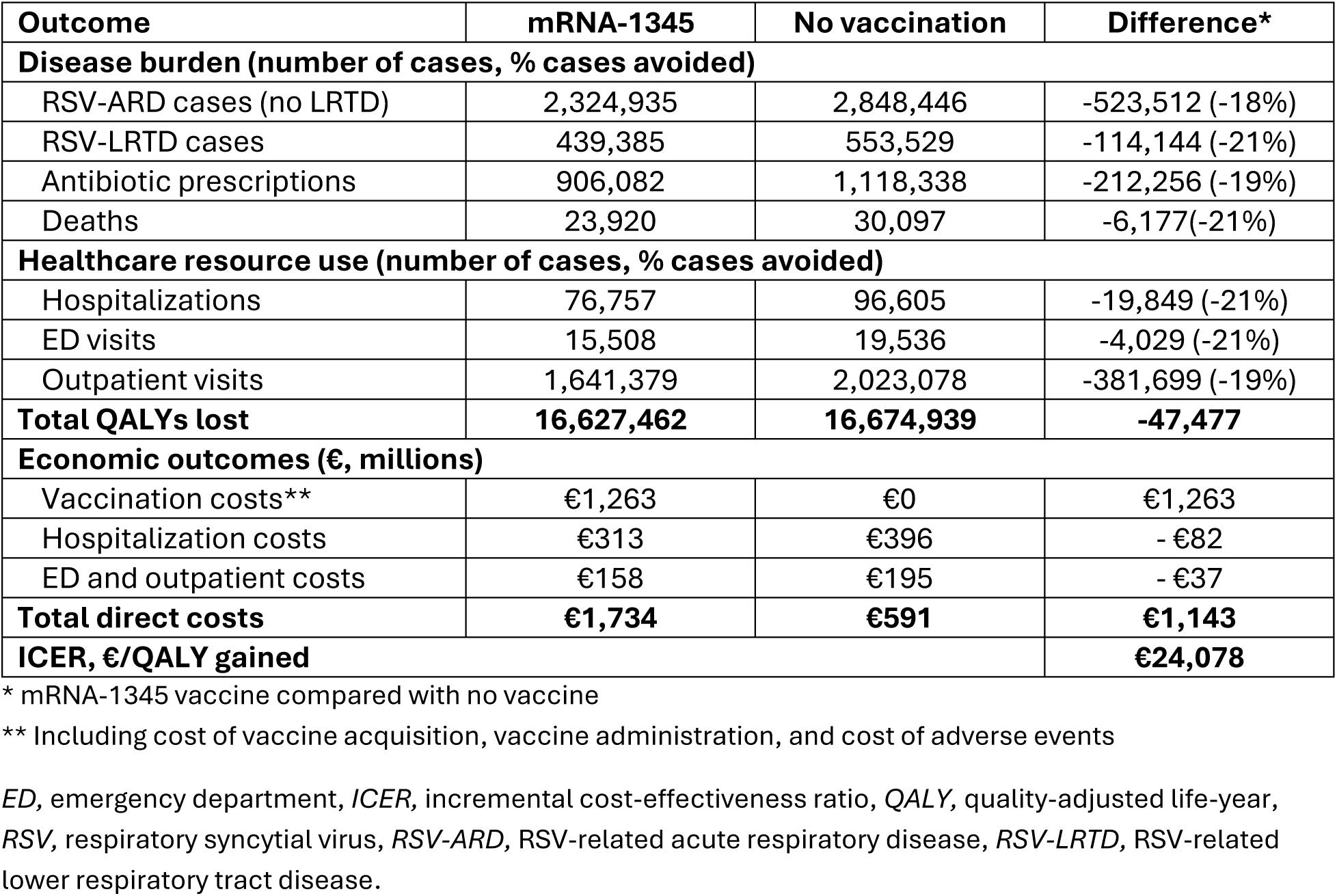
Base-case analysis results, over 5 years ED, emergency department, ICER, incremental cost-effectiveness ratio, QALY, quality-adjusted life-year, RSV, respiratory syncytial virus, RSV-ARD, RSV-related acute respiratory disease, RSV-LRTD, RSV-related lower respiratory tract disease.

Vaccination with mRNA-1345 in the target population was estimated to cost over €1 billion, including vaccine administration costs, acquisition costs, and costs related to adverse events. Cost savings due to avoided healthcare resource use were estimated at €120 million. Overall, the total incremental cost of €1,143 million and the additional 47,477 QALYs gained resulted in an ICER of €24,078 (Table 3). This is lower than the commonly referenced WTP range of €33,000-40,000 per QALY gained, indicating that the mRNA-1345 strategy is a cost-effective approach.

### Scenario Analysis

The results of the scenario analyses comparing mRNA-1345 to no vaccine are presented in Table 4 in comparison to the base-case results, with hospitalizations avoided and ICER provided as key outcomes. Detailed results for each scenario are presented in Supplementary Materials.

**Table 4.**
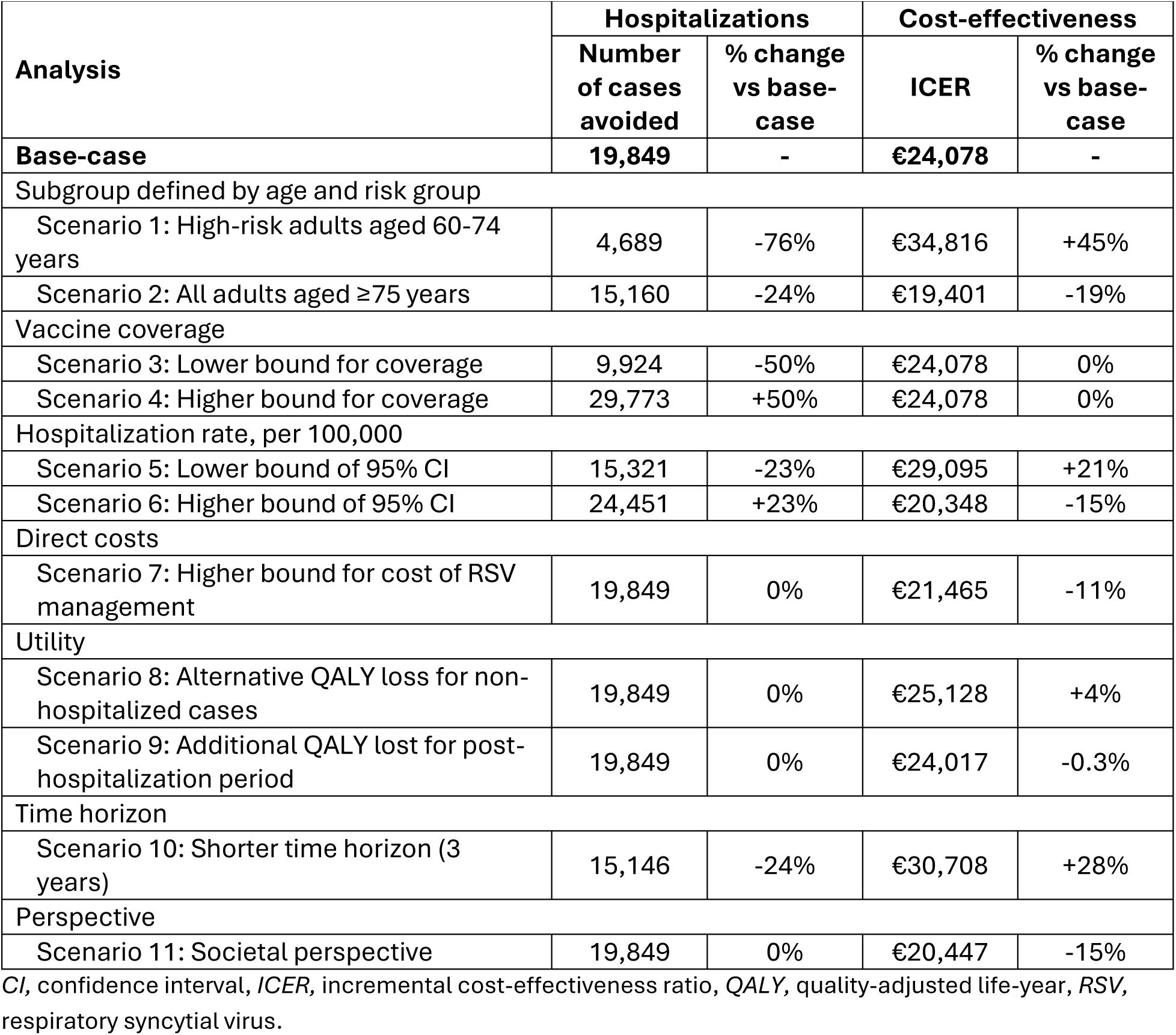
Scenario analysis results CI, confidence interval, ICER, incremental cost-effectiveness ratio, QALY, quality-adjusted life-year, RSV, respiratory syncytial virus.

| Analysis | Hospitalizations |  | Cost-effectiveness |  |
| --- | --- | --- | --- | --- |
|  | Number of cases avoided | % change vs base-case | ICER | % change vs base-case |
| **Base-case** | **19,849** | **-** | **€24,078** | **-** |
| Subgroup defined by age and risk group |  |  |  |  |
| Scenario 1: High-risk adults aged 60-74 years | 4,689 | -76% | €34,816 | +45% |
| Scenario 2: All adults aged ≥75 years | 15,160 | -24% | €19,401 | -19% |
| Vaccine coverage |  |  |  |  |
| Scenario 3: Lower bound for coverage | 9,924 | -50% | €24,078 | 0% |
| Scenario 4: Higher bound for coverage | 29,773 | +50% | €24,078 | 0% |
| Hospitalization rate, per 100,000 |  |  |  |  |
| Scenario 5: Lower bound of 95% CI | 15,321 | -23% | €29,095 | +21% |
| Scenario 6: Higher bound of 95% CI | 24,451 | +23% | €20,348 | -15% |
| Direct costs |  |  |  |  |
| Scenario 7: Higher bound for cost of RSV management | 19,849 | 0% | €21,465 | -11% |
| Utility |  |  |  |  |
| Scenario 8: Alternative QALY loss for non-hospitalized cases | 19,849 | 0% | €25,128 | +4% |
| Scenario 9: Additional QALY lost for post-hospitalization period | 19,849 | 0% | €24,017 | -0.3% |
| Time horizon |  |  |  |  |
| Scenario 10: Shorter time horizon (3 years) | 15,146 | -24% | €30,708 | +28% |
| Perspective |  |  |  |  |
| Scenario 11: Societal perspective | 19,849 | 0% | €20,447 | -15% |

Overall, in ten out of eleven scenarios, vaccination with mRNA-1345 was shown to be cost-effective compared with no vaccination, with ICERs below the commonly referenced WTP value of €33,000 per QALY gained. In one scenario, ICER slightly exceeded €33,000 per QALY gained but remained below the higher indicative WTP value of €40,000 per QALY gained. These values are used as reference points in the absence of an officially established WTP threshold in Italy.

Scenarios 1 and 2 explored the impact of vaccination in the two target subgroups separately. Restricting vaccination to high-risk adults aged 60-74 years resulted in an ICER of €34,816 per QALY gained, marginally exceeding the commonly referenced WTP value of €33,000 per QALY gained. Nevertheless, vaccination in this subgroup was still estimated to prevent more than 4,600 RSV-related hospitalizations, indicating a substantial public health impact. Vaccination of all adults aged ≥75 years remained cost-effective, with an ICER of €19,401 per QALY gained. More than 15,100 RSV-related hospitalizations were prevented over the 5-year time horizon, highlighting the considerable preventable burden of severe RSV disease among adults of advanced age.

Scenarios 3 and 4 explored alternative coverage assumptions. In these scenarios, no changes to ICER are expected, as economic and health outcomes of the vaccination campaign change linearly accordingly to the uptake, i.e., the same incremental benefits are expected for each additional part of the population receiving the vaccine, and the efficiency of the program remains stable. However, these scenarios demonstrated the range of the hospitalizations avoided, which can be expected depending on the achieved coverage, with a considerable number of hospitalizations avoided in both scenarios (more than 9,900 and 29,700 hospitalizations avoided with coverage of 25% and 75%, respectively).

Scenarios 5 and 6 explored uncertainty in the underlying RSV burden by applying the lower and upper 95% CI bounds of the estimated hospitalization rate reported by Osei-Yeboah et al. (4). As expected, the projected public health impact varied in line with the underlying RSV burden, while the conclusion regarding cost-effectiveness remained unchanged.

In Scenarios 7-9, which considered alternative data sources for key cost and utility inputs, no substantial changes to the results were observed, supporting the robustness of the base-case input selection.

Scenario 10 applied a conservative 3-year time horizon, limiting the duration over which vaccine benefits were accrued. Despite preventing fewer RSV-related hospitalizations due to the omitted vaccination benefits for years 4 and 5, vaccination remained cost-effective, with an ICER of €30,708 per QALY gained.

Finally, in Scenario 11, which explored the societal perspective, a relatively minor decrease in the ICER was projected. This indicates that RSV prevention in older adults has the potential to reduce productivity losses, which might become more important in the future, considering the population aging trend and an increasing proportion of individuals in this age group which remains economically active.

### Sensitivity Analyses

Results of the DSA are presented in Figure 2. Among all included parameters, the percentage of individuals experiencing RSV-ARD, RSV-LRTD, percentage requiring hospitalization, and percentage of hospitalized cases that result in death had the highest impact on the ICER. Other noted impactful parameters were QALY loss per RSV case and discount rate for QALYs. Variation of all the remaining inputs change the ICER by less than 5%. Across all DSA simulations, ICERs were between €18,564 and €32,172, remaining below the commonly referenced WTP range of €33,000-40,000 per QALY gained.

**Figure 2.**
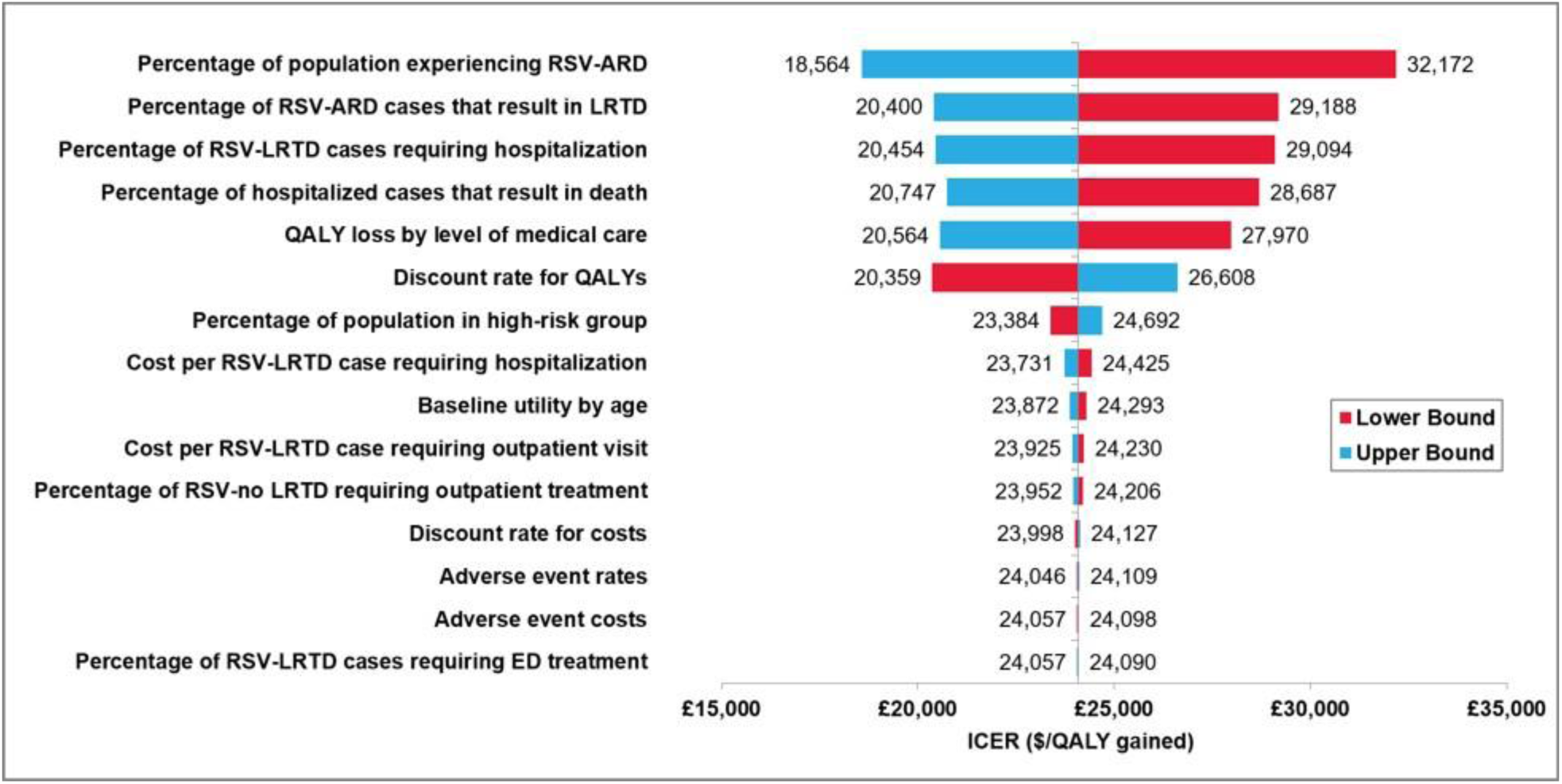
Deterministic sensitivity analysis results: Tornado Diagram ED, emergency department, ICER, incremental cost-effectiveness ratio, QALY, quality-adjusted life-year, RSV, respiratory syncytial virus, RSV-ARD, RSV-related acute respiratory disease, RSV-LRTD, RSV-related lower respiratory tract disease.

Results of the PSA are presented in Figure 3. For the mRNA-1345 strategy, the mean QALY gain was 47,585 (95% CI: 29,724, 70,361), and the mean incremental costs were €1,137 million (95% CI: €896 million, €1,403 million), resulting in a probabilistic ICER of €23,889 per QALY gained. This ICER is below the reference WTP values. The probability that the mRNA-1345 vaccination strategy is cost-effective compared to no vaccination was 93% and 99% at the WTP values of €33,000 and €40,000 per QALY gained, respectively. The CEAC indicates that this probability exceeds 50% at WTP values above €24,367 per QALY gained.

**Figure 3.**
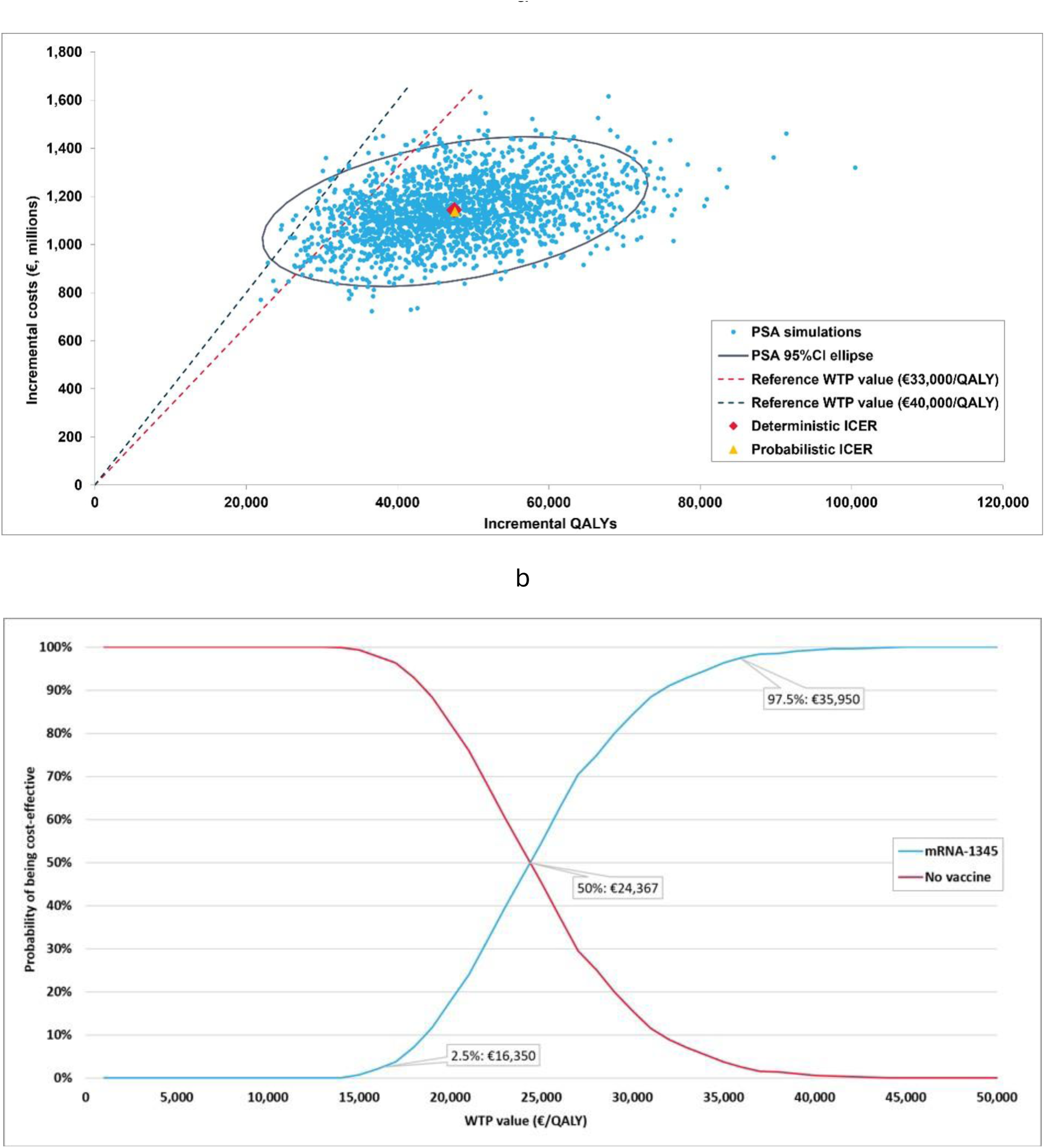
Probabilistic sensitivity analysis results: a. Cost-effectiveness plane. b. Cost-effectiveness acceptability curve. CI, confidence interval, ICER, incremental cost-effectiveness ratio, PSA, probabilistic sensitivity analyses, QALY, quality-adjusted life year, WTP, willingness-to-pay.

## Discussion

Prevention of RSV-related diseases is increasingly recognized as a public health priority in Italy, particularly among older and high-risk adults, reflecting the substantial burden of RSV in this population, the availability of effective preventive interventions, and the recent introduction of adult RSV vaccination programs in several European countries. Current recommendations from Italian scientific societies prioritize RSV vaccination for adults aged 60-74 years with chronic medical conditions and all adults aged ≥75 years, while nation-wide reimbursement and implementation decisions for adult RSV vaccination are currently under evaluation (1, 19).

The present analysis supports these recommendations by demonstrating that vaccination with mRNA-1345 has the potential to substantially reduce the public health burden of RSV by preventing 21% of hospitalizations, 21% of ED visits and 19% of outpatient visits associated with RSV. Beyond the direct clinical benefits, the projected decrease in healthcare utilization could help alleviate pressure on healthcare services during RSV season, particularly in the context of population aging and increasing healthcare demand in Italy (69). In addition, the projected 19% reduction in antibiotic prescribing represents a further potential benefit by reducing pharmaceutical expenditure, while supporting antimicrobial stewardship efforts and potentially limiting the development of antimicrobial resistance. From an economic perspective, the projected health benefits were associated with a favorable cost-effectiveness profile. The base-case and probabilistic ICERs were estimated at €24,078 and €23,889, respectively, below the commonly referenced WTP range of €33,000-€40,000 per QALY gained. Sensitivity analyses have shown that the analysis results were robust to changes in all of the model parameters, with the ICER remaining below the WTP value of €33,000 across all DSA simulations and in ten out of eleven explored scenarios. In one scenario, the ICER slightly exceeded €33,000, but remained below the alternative WTP value of €40,000 per QALY gained. The percentage of PSA iterations demonstrating cost-effectiveness was estimated at 93% and 99% for the WTP values of €33,000 and €40,000 per QALY gained, respectively.

To date, four economic evaluations of RSV vaccination in older adults have been published for the Italian setting (20–23). Puggina et al. (22) reported that vaccination with adjuvanted RSVPreF3 was cost-effective in adults aged 60-74 years with chronic obstructive pulmonary disease, with a subsequent analyses by Calabrò et al. (20) and Puggina et al. (23) demonstrating cost-effectiveness in a broader population of older adults, including those at increased risk. Likewise, Di Matteo et al. (21) reported that bivalent prefusion F vaccination was cost-effective in adults aged 60-74 years with comorbidities as well as all adults aged ≥75 years. Direct comparison of ICER estimates across studies is not appropriate, due to differences in the modeling approach, vaccine-specific effectiveness assumptions derived from separate clinical trials in the absence of head-to-head comparative evidence, and the Italy-specific RSV burden estimates used to populate the models, which remain uncertain. Nevertheless, all available Italian studies consistently demonstrate that RSV vaccination is a cost-effective intervention in older adults, indicating that its implementation at a national level would be an efficient use of Italian National Health Service resources.

This study has several strengths and limitations that should be considered.

A key strength of this study is the use of the most up-to-date Italy-specific epidemiological, demographic and healthcare resource use data wherever available, together with clinical and economic inputs relevant to the Italian National Health Service. In particular, the present analysis relies on the model calibrated to age-specific hospitalization rate estimates reported by Osei-Yeboah et al. (4). This study was selected to inform a calibration target, as it provides the first harmonized assessment of RSV-associated hospitalization burden in adults across the European Union and has been cited by recent Italian expert reviews as a relevant source for estimating the burden of RSV in the country (1, 19). Additionally, RSV-related mortality inputs considered both in-hospital and one-month post-discharge deaths, informed by a recent Italian database study by Puggina et al. (2), thereby capturing emerging evidence on consequences of severe RSV in older adults beyond in-hospital outcomes. While remaining uncertain due to data limitations, this approach is expected to be conservative, as it does not consider the impact of multimorbidity, worsening of underlying diseases, re-hospitalization, or the increased long-term risk of death, beyond one-month post-discharge mortality risk (2).

This study is subject to several limitations, primarily driven by scarcity of data informing the underlying epidemiological and vaccine-related inputs. Overall, data on the true burden of RSV in older adults in Italy remain limited, and the available estimates remain substantially under-ascertained (7, 70). Therefore, where Italy-specific evidence was unavailable, selected epidemiological inputs were informed by studies conducted in other high-income countries, including the probability of RSV-ARD (31), the probability of hospitalization following RSV-LRTD (33), the relative risk of hospitalization among adults with chronic medical conditions (34), and the under-ascertainment adjustment factor (11). Another limitation of this analysis is that vaccine efficacy inputs were derived from clinical trial data with follow-up limited to 24 months. As such, extrapolation of protection beyond this period is uncertain. However, a scenario analysis which applied a shorter 3-year time horizon confirmed the robustness of the conclusion regarding cost-effectiveness. It should be also noted that RSV incidence varies year to year (12), therefore estimate of vaccine economic value depends in part on the severity of the season, as reported for influenza vaccination (71, 72).

Several conservative assumptions and alternative input values were preferentially adopted to avoid overestimating the clinical and economic value of vaccination. Importantly, the increased risk of severe RSV outcomes in individuals with chronic medical conditions was captured only for a higher probability of hospitalization following RSV-LRTD. In the absence of robust data, this analysis did not account for the broader clinical and economic consequences of RSV among older adults with chronic medical conditions, including differences in disease severity, healthcare resource use and costs, caregiver burden, worsening of underlying diseases, re-hospitalization, and increased long-term mortality beyond one month after discharge (2). The model also did not account for the potentially greater burden of RSV among older adults with multimorbidity. In addition, it was assumed that the target population is at risk of only one infection per season; however, if individuals acquired more than one infection per season and both can be prevented by the vaccine, the value of the vaccine would increase. Vaccine was assumed to impact hospitalization and LRTD rates equally, but more than RSV-ARD incidence. However, emerging evidence suggests that vaccines provide much greater protection against hospitalization compared to less severe outcomes (e.g., RSV-ARD or RSV-LRTD), and therefore, our analysis is expected to be conservative relative to the real-world vaccine effectiveness (28, 73). Finally, this analysis was conducted using a cohort-level static model, which does not capture indirect protection or health benefits for new cohorts becoming eligible for vaccination over time, therefore, the overall public health impact of RSV vaccination program is likely to be underestimated.

Despite recent advances in understanding RSV epidemiology, important evidence gaps remain. Emerging evidence suggests that the burden of RSV extends beyond acute infection, with severe disease potentially contributing to the worsening of underlying health conditions, re-hospitalization and long-term risk of excess mortality following hospitalization in vulnerable older adults (2). A better understanding of these longer-term consequences, including the extent to which vaccination may mitigate them, as well as broader benefits of RSV prevention for patients, caregivers, and society, would allow future economic evaluations to better capture the full value of RSV vaccination. Continued evidence generation on the burden of RSV, long-term vaccine effectiveness, and duration of protection will also enable optimization of adult RSV vaccination strategies, including decisions regarding the eligibility criteria and revaccination policy.

## Conclusion

Vaccination with mRNA-1345 represents a cost-effective strategy for the prevention of RSV disease among high-risk adults aged 60-74 years and all adults aged ≥75 years in Italy. By preventing thousands of hospitalizations and deaths, vaccination has the potential to substantially reduce the clinical and healthcare burden of RSV in these populations while providing good value for the Italian National Health Service. These findings provide robust economic and public health evidence to inform national immunization policy and reimbursement decision-making and support the implementation of RSV vaccination among older adults in Italy.

## Data Availability

All data generated are available in the electronic supplementary materials accompanying this manuscript.

## Acknowledgements

Medical Writing, Editorial and Other Assistance

No medical writing, editorial, or other assistance was provided for the preparation of this manuscript.

## Author contributions

Mariia Dronova, Camille Moyon, Lukasz Pyrek, Katherine Hicks, Ziyi Xiao, Filippo Rumi, Chiara de Waure, Stefan Scholz and Parinaz Ghaswalla contributed to the study conception and design. The model was developed by Katherine Hicks and Ziyi Xiao. Data collection, analysis, interpretation of results, and development of the first draft of the manuscript were performed by Mariia Dronova, Camille Moyon and Lukasz Pyrek. All authors commented on previous versions of the manuscript. All authors read and approved the final manuscript.

## Data Availability

Data sources for this study are publicly available, as described in Table 1 and Supplementary materials.

## Open Access

This article is licensed under a Creative Commons Attribution-NonCommercial 4.0 International License, which permits any non-commercial use, sharing, adaptation, distribution and reproduction in any medium or format, as long as you give appropriate credit to the original author(s) and the source, provide a link to the Creative Commons license, and indicate if changes were made. The images or other third party material in this article are included in the article’s Creative Commons license, unless indicated otherwise in a credit line to the material. If material is not included in the article’s Creative Commons license and your intended use is not permitted by statutory regulation or exceeds the permitted use, you will need to obtain permission directly from the copyright holder. To view a copy of this license, visit http://creativecommons.org/licen <u>ses/by-nc/4.0/</u>.

