## Supplement for "Cost-Effectiveness Analysis of the mRNA-1345 RSV Vaccine for Older Adults in Italy"

*Paris, France*

**

### SUPPLEMENTARY METHODS

#### Methods For Estimating Inputs That Determine RSV Outcomes In The Decision Tree

This section outlines the methods used to estimate the following model inputs:

- Percentage of RSV-related acute respiratory disease cases (RSV-ARD) that result in lower respiratory tract disease (LRTD)
- Percentage of RSV-related LRTD cases that result in hospitalization
- Percentage of RSV-related LRTD cases requiring hospitalization that result in death

The applied stepwise procedure is described below.

**Step 1. Estimation of hospitalization rates in the general population (calibration target).**

The age-specific RSV hospitalization incidence reported for adults in Italy by Osei-Yeboah et al. was considered representative of the general Italian population (1). The study reported Italy-specific incidence for the following age groups: 18-64, 65-74 years and 85 years and older. However, it could be expected that risk of severe outcomes increases with age. Using these data for calibration directly could lead to underestimation of hospitalization rate in 60-64 year-olds. Therefore, the hospitalization rate for the 60-64-year age group was estimated as the average between the estimates reported for adults aged 18-64 years and 65-74 years. The estimates reported for 65-74 year-olds, 75-84 year-olds, and ≥85 year-olds were applied directly. The values used as the calibration target are provided in Table S1.

*Table S1. Calibration target: Hospitalization rate per 100,000, general population*

| Age group | Hospitalization rate per 100,000, general population | Source |
|---|---|---|
| 60-64 years | 30* | Osei-Yeboah et al. (1) |
| 65-69 years | 55 | |
| 70-74 years | 55 | |
| 75-79 years | 221 | |
| 80-84 years | 221 | |
| ≥85 years | 231 | |

* In the absence of age-specific data for adults aged 60-64 years, the value was estimated as the average of the estimates reported for adults aged 18-64 years and 65-74 years. Reported hospitalization rates for adults aged 18-64 years and 65-74 years were 4 and 55 per 100,000, respectively (1).

**Step 2. Estimation of percentage of RSV-related LRTD cases that result in hospitalization.**

Calibration also required an estimate of the percentage of patients with RSV-LRTD requiring hospitalization. The data for the general population were derived from Fleming et al. (2). To account for under-ascertainment, the percentage of RSV-LRTD patients requiring hospitalization was adjusted using an under-ascertainment adjustment factor of 1.5, informed by McLaughlin et al. (3). Further, to account for an increased risk of hospitalization in patients with chronic medical conditions, a conservative risk ratio (RR) of 1.5 was applied. This value was sourced from the modeling study by Osei-Yeboah et al. (4). This study reports RRs for high-risk versus general population ≥45 years in the UK and Denmark, stratified by the underlying medical condition (dataset 1), and stratified by both the underlying medical condition and age (dataset 2). Only dataset 1 was used for this analysis, as it was not clear whether age-specific estimates accurately reflect the trends observed in clinical practice (e.g., the reported RR for some age groups were above 6.0, and the age trends were inconsistent or counterintuitive). Then, as a conservative assumption, the lowest reported value from the dataset 1 was applied for this analysis. This RR of 1.5 was applied only to individuals aged 60-74 years, assuming that the reported hospitalization rate for the age group of ≥75 years already captured the combined risk due to age and chronic medical conditions.

The hospitalization rate of the general population was used to derive the hospitalization rate for the high-risk population using the following formula:

$$ARgp = \%POPhr * ARhr + \%POPlr * ARlr,$$

where: AR is absolute risk; %POP is percentage of population; gp is general population; hr is high risk; lr is low risk (here and in the formulas below).

Replacing ARhr with ARgp * RR (where RR is the relative risk for the high-risk population compared to the general population), we get:

$$ARgp = \%POPhr * ARgp * RR + \%POPlr * ARlr$$

This formula has been rearranged to derive the hospitalization rate in the LR population ($AR_{LR}$):

$$ARlr = (ARgp - (\%POPhr * ARgp * RR))/\%POPlr$$

The hospitalization rate for the HR population was then calculated as:

$$ARhr = ARgp * RR$$

The estimated data are summarized in Table S2.

*Table S2. Calculation of percentage of RSV-related LRTD cases that result in hospitalization*

| **Age group** | **Relative risk (high risk vs general population)** | **Percentage of RSV-related LRTD cases that result in hospitalization** | | | | **Source** |
|---|---|---|---|---|---|---|
| | | **General population, unadjusted** | **General population, adjusted by underreporting factor of 1.5** | **Non-high risk** | **High-risk** | |
| 60-64 years | 1.5 | 3.9% | 5.9% | 3.1% | 8.8% | Fleming et al. (2), McLaughlin et al. (3), Osei-Yeboah et al. (4). |
| 65-69 years | 1.5 | 6.8% | 10.1% | 5.3% | 15.2% | |
| 70-74 years | 1.5 | 6.8% | 10.1% | 5.3% | 15.2% | |
| 75-79 years | 1 | 12.4% | 18.7% | 18.7% | 18.7% | |
| 80-84 years | 1 | 12.4% | 18.7% | 18.7% | 18.7% | |
| ≥85 years | 1 | 12.4% | 18.7% | 18.7% | 18.7% | |

*RSV,* respiratory syncytial virus, *RSV-LRTD,* RSV-related lower respiratory tract disease.

**Step 3. Calibration of the percentage of RSV-ARD patients who develop RSV-LRTD.**

The percentage of RSV-ARD cases that result in LRTD was calibrated to match the age-specific RSV hospitalization rates in the general population. The following data points were used as calibration inputs: RSV-ARD incidence rate (described in the main manuscript), percentage of RSV-LRTD patients requiring hospitalization and the general population RSV hospitalization rate (described in steps above).

For each age group, the RSV hospitalization rate can be calculated as:

$$HospRate_i = ARD_i \times LRTD\_ARD_i \times Hosp\_LRTD_i$$

where: $HospRate_i$ is the RSV hospitalization rate, $ARD_i$ is the RSV-ARD incidence rate, $LRTD\_ARD_i$ is the % of RSV-ARD patients who develop RSV-LRTD, and i denotes the age group.

Solving for $LRTD\_ARD_i$ , the calibrated model input was calculated as:

$$LRTD\_ARD_i = \frac{HospRate_i}{ARD_i \times Hosp\_LRTD_i}$$

The calibrated values were applied as model inputs, for high-risk and non-high risk individuals. The input values are provided in Table S3.

*Table S3. Calibrated percentage of RSV-ARD cases that result in LRTD*

| **Age group** | **Percentage of RSV-ARD cases that result in LRTD** |
|---|---|
| 60-64 years | 8.8% |
| 65-69 years | 9.6% |
| 70-74 years | 9.6% |
| 75-79 years | 20.9% |
| 80-84 years | 20.9% |
| ≥85 years | 21.8% |

*LRTD,* lower respiratory tract disease, *RSV,* respiratory syncytial virus, *RSV-ARD,* RSV-related acute respiratory disease.

#### Estimation of RSV-Attributable Mortality

The percentage of hospitalized RSV-LRTD cases resulting in death was estimated using the combined in-hospital and 1-month post-discharge mortality data reported by Puggina et al. (5), which was adjusted to account for all-cause mortality (6).

The applied stepwise procedure is described below.

**Step 1.** The reported numbers of hospitalized patients, in-hospital deaths, and deaths within 1 month after hospitalization were extracted from Puggina et al. (5) for each reported age category (≥60, ≥65, and ≥75 years).

**Step 2.** The extracted cumulative estimates were recomputed into the age-specific estimates for the modeled age groups (60-64, 65-74, and ≥75 years), and the corresponding raw probabilities of in-hospital death and cumulative 1-month mortality following hospitalization were calculated.

**Step 3.** Probability of death within 1 month after hospitalization was converted to a hazard. To avoid double-counting, the hazard of 1-month all-cause mortality (informed by ISTAT (6)) was then subtracted from the cumulative 1-month mortality hazard to estimate a 1-month hazard of RSV-attributable post-discharge mortality. The obtained adjusted hazard was converted back to probability.

In-hospital death was considered to occur within a short timeframe of the hospital stay, therefore probability-to-hazard conversion and adjustment for all-cause mortality was not required for this outcome (6).

**Step 4.** The overall probability of death following hospitalization was calculated as the cumulative probability of in-hospital death and RSV-attributable 1-month post-discharge death, using the following formula:

$$P(total\ death) = P(in - hospital) + (1 - P(in - hospital)) \times P(post - discharge)$$

#### Projected Vaccine Effectiveness Curves

The nonlinear models were fitted to monthly trial data. The model used the logarithm of time or log(t) as the independent variable and monthly log incidence rate ratio or log(IRR) as the dependent variable to fit the following curve:

$$VE(t) = 1 - exp(b0 + b1 \times \log[t])$$

Inside the exponential, we modeled the log-IRR as a linear function of log(t):

$$exp(b0 + b1 * \log[t]) = IRR\ at\ time\ t$$

Vaccine effectiveness was then derived as 1-IRR. Using log(t) allows for a flexible nonlinear waning pattern, which avoids the assumption that the IRR decays linearly in absolute time. Instead, the effect is smoother and often fits real-world waning curves better, since the strongest changes are usually seen early on, then taper off.

The fitted equations used to project vaccine efficacy waning for each outcome are presented below.

$$VE\ RSV\_ARD(t) = 1 - exp(-2.5107449 + 0.3057026 \times \log[t])$$

$$VE\ RSV\_LRTD(t) = 1 - exp(-1.9787039 + 0.2344412 \times \log[t])$$

### SUPPLEMENTARY TABLES

*Table S4. Base-Case Inputs, DSA Ranges, And Assigned PSA Parameters And Distributions*

| Model input parameter | Base-case value | Source of base-case value range | DSA range | Source of DSA range | PSA parameters and distribution | Source of distribution parameter(s) |
|---|---|---|---|---|---|---|
| **Settings** | | | | | | |
| Discount rates for costs and outcomes | 3.0% | AIFA HTA Guidelines (7) | 0.0% – 5.0% | AIFA HTA Guidelines (7) | Not varied | – |
| Time horizon (in years) | 5 | ISPOR-SMDM Modeling Good Research Practices Task Force (8) | Not varied | – | Not varied | – |
| **Population** | | | | | | |
| Italian population size, by age group | | | | | | |
| 60-64 years | 4,566,052 | Italian National Institute of Statistics (ISTAT) (9) | Not varied | – | Not varied | – |
| 65-69 years | 3,831,068 | | | | | |
| 70-74 years | 3,278,863 | | | | | |
| 75-79 years | 3,095,916 | | | | | |
| 80-84 years | 2,103,261 | | | | | |
| 85+ years | 2,511,392 | | | | | |
| Percentage of male in the target population | 45.4% | Italian National Institute of Statistics (ISTAT) (9) | Not varied | – | Not varied | – |
| Percentage of individuals at high risk, by age group* | | | | | | |
| 60-64 years | 48.9% | PASSI d'Argento surveillance system (10), assuming the | 39.1% – 58.7% | ±20% of base-case value | Beta (α = 48.37; β = 50.55) | Calibrated to the assumed +/-20% range** |
| 65-69 years | 48.9% | | 39.1% – 58.7% | | Beta (α = 48.37; β = 50.55) | |

| Model input parameter | Base-case value | Source of base-case value range | DSA range | Source of DSA range | PSA parameters and distribution | Source of distribution parameter(s) |
|---|---|---|---|---|---|---|
| 70-74 years | 48.9% | same percentage for individuals aged 60-64 years as reported for those aged 65-74 years | 39.1% – 58.7% | | Beta (α = 48.37; β = 50.55) | |
| 75-79 years | 62.5% | | 50.0% – 75.0% | | Beta (α = 35.25; β = 21.15) | |
| 80-84 years | 62.5% | | 50.0% – 75.0% | | Beta (α = 35.25; β = 21.15) | |
| 85+ years | 71.6% | | 57.3% – 85.9% | | Beta (α = 26.80; β = 10.63) | |
| **Vaccination coverage rate and effectiveness** | | | | | | |
| mRNA-1345 vaccine coverage, all age and risk groups | 50.0% | Assumption based on coverage reported for influenza vaccination in 2024-2025 season (11) | Not varied | – | Beta (α = 47.30; β = 47.30) | Calibrated to the assumed +/-20% range** |
| Estimated average annual vaccine efficacy against RSV-ARD | | | | | | |
| Initial VE, 1 month post-vaccination | 69.2% | Evidence from clinical trial data (12), projection over 5 years | Not varied | – | Not varied | – |
| Year 1 post-vaccination | 53.9% | | | | | |
| Year 2 post-vaccination | 39.2% | | | | | |
| Year 3 post-vaccination | 31.5% | | | | | |
| Year 4 post-vaccination | 25.9% | | | | | |
| Year 5 post-vaccination | 21.4% | | | | | |
| Estimated average annual vaccine efficacy against RSV-LRTD (hospitalized and non-hospitalized cases) | | | | | | |
| Initial VE, 1 month post-vaccination | 76.9% | | Not varied | – | Not varied | – |

| Model input parameter | Base-case value | Source of base-case value range | DSA range | Source of DSA range | PSA parameters and distribution | Source of distribution parameter(s) |
|---|---|---|---|---|---|---|
| Year 1 post-vaccination | 60.7% | Evidence from clinical trial data (12), projection over 5 years | | | | |
| Year 2 post-vaccination | 43.9% | | | | | |
| Year 3 post-vaccination | 34.5% | | | | | |
| Year 4 post-vaccination | 27.5% | | | | | |
| Year 5 post-vaccination | 21.7% | | | | | |
| **Vaccine-related adverse events, all age and risk groups** | | | | | | |
| Adverse event rates, per 100,000 doses | | | | | | |
| Grade 3 local | 1,426 | Wilson et al. (13) | 1,141 – 1,711 | ±20% of base-case value | Gamma (α = 96.79; θ = 14.73) | Calibrated to the assumed +/-20% range** |
| Grade 3 systemic | 1,133 | | 906 – 1,360 | | Gamma (α = 96.79; θ = 11.71) | |
| Adverse event costs | | | | | | |
| Grade 3 local | €28.56 | AIFA transparency lists (14), Capri et al. (15), ISTAT (16, 17) | €22.85 – €34.27 | ±20% of base-case value | Gamma (α = 96.79; θ = 0.30) | Calibrated to the assumed +/-20% range** |
| Grade 3 systemic | €28.53 | AIFA transparency lists (14), Ministry of Health decree of November 25, 2024 (18), ISTAT (16) | €22.82 – €34.24 | | Gamma (α = 96.79; θ = 0.29) | |
| QALY loss per adverse event | | | | | | |
| Grade 3 local | 0.0003 | Estimated based on Prosser et al. (19) | 0.00022 – 0.00033 | ±20% of base-case value | Beta (α = 384.04; β = 1,402,323.65) | Calibrated to the assumed +/-20% range** |
| Grade 3 systemic | 0.0011 | | 0.00088 – 0.00131 | | Beta (α = 383.72; β = 350,004.34) | |

| Model input parameter | Base-case value | Source of base-case value range | DSA range | Source of DSA range | PSA parameters and distribution | Source of distribution parameter(s) |
|---|---|---|---|---|---|---|
| **RSV epidemiology and clinical outcomes (without vaccination)** | | | | | | |
| RSV seasonality by month | | | | | | |
| October 2023 | 5.3% | EpiCentro, 2023-2024 season (20) | Not varied | – | Not varied | – |
| November 2023 | 16.4% | | | | | |
| December 2023 | 29.7% | | | | | |
| January 2024 | 23.4% | | | | | |
| February 2024 | 11.1% | | | | | |
| March 2024 | 8.4% | | | | | |
| April 2024 | 5.9% | | | | | |
| May 2024 | 0.0% | | | | | |
| June 2024 | 0.0% | | | | | |
| July 2024 | 0.0% | | | | | |
| August 2024 | 0.0% | | | | | |
| September 2024 | 0.0% | | | | | |
| Percentage experiencing RSV-ARD, all age and risk groups | 5.7% | Korsten et al. (21) | 4.4% – 7.2% | 95% CI estimated from Korsten et al. (21) | Beta (α = 59; β = 981) | Korsten et al. (21) |
| Percentage experiencing RSV-LRTD, by age group | | | | | | |
| 60-64 years | 8.8% | Estimated based on: Korsten et al. (21), Fleming et al. (2), McLaughlin et al. (3) and Osei-Yeboah et al. (1) | 7.1% – 10.6% | ±20% of base-case value | Beta (α = 87.99; β = 907.33) | Calibrated to the assumed +/-20% range** |
| 65-69 years | 9.6% | | 7.6% – 11.5% | | Beta (α = 87.28; β = 826.42) | |
| 70-74 years | 9.6% | | 7.6% – 11.5% | | Beta (α = 87.28; β = 826.42) | |
| 75-79 years | 20.9% | | 16.7% – 25.1% | | Beta (α = 76.02; β = 288.12) | |
| 80-84 years | 20.9% | | 16.7% – 25.1% | | Beta (α = 76.02; β = 288.12) | |
| 85+ years | 21.8% | | 17.5% – 26.2% | | Beta (α = 74.86; β = 268.21) | |

| Model input parameter | Base-case value | Source of base-case value range | DSA range | Source of DSA range | PSA parameters and distribution | Source of distribution parameter(s) |
|---|---|---|---|---|---|---|
| Percentage of RSV-LRTD patients requiring hospitalization, non-high-risk individuals, by age group | | | | | | |
| 60-64 years | 3.1% | Estimated based on: Fleming et al. (2), McLaughlin et al. (3), Osei-Yeboah et al. (4), and Italian PASSI d'Argento surveillance system (10) | 2.5% – 3.7% | ±20% of base-case value | Beta (α = 93.06; β = 2,940.37) | Calibrated to the assumed +/-20% range** |
| 65-69 years | 5.3% | | 4.2% – 6.4% | | Beta (α = 91.52; β = 1,637.44) | |
| 70-74 years | 5.3% | | 4.2% – 6.4% | | Beta (α = 91.52; β = 1,637.44) | |
| 75-79 years | 18.7% | | 14.9% – 22.4% | | Beta (α = 78.22; β = 340.96) | |
| 80-84 years | 18.7% | | 14.9% – 22.4% | | Beta (α = 78.22; β = 340.96) | |
| 85+ years | 18.7% | | 14.9% – 22.4% | | Beta (α = 78.22; β = 340.96) | |
| Percentage of RSV-LRTD patients requiring hospitalization, high-risk individuals, by age group | | | | | | |
| 60-64 years | 8.8% | Estimated based on: Fleming et al. (2), McLaughlin et al. (3), and Osei-Yeboah et al. (4) | 7.1% – 10.6% | ±20% of base-case value | Beta (α = 88.00; β = 909.36) | Calibrated to the assumed +/-20% range** |
| 65-69 years | 15.2% | | 12.2% – 18.3% | | Beta (α = 81.63; β = 454.58) | |
| 70-74 years | 15.2% | | 12.2% – 18.3% | | Beta (α = 81.63; β = 454.58) | |
| 75-79 years | 18.7% | | 14.9% – 22.4% | | Beta (α = 78.22; β = 340.96) | |
| 80-84 years | 18.7% | | 14.9% – 22.4% | | Beta (α = 78.22; β = 340.96) | |
| 85+ years | 18.7% | | 14.9% – 22.4% | | Beta (α = 78.22; β = 340.96) | |
| Percentage of RSV-LRTD patients requiring ED treatment, all age and risk groups | 3.5% | Baglivo et al. (22) | 0.7% – 8.3% | 95% CI estimated from Baglivo et al. (22) | Beta (α = 3.00; β = 82.00) | Baglivo et al. (22) |
| Percentage of RSV-No LRTD patients requiring | 55.7% | Dal Negro et al. (23) | 51.9% – 59.4% | 95% CI estimated from Dal Negro et al. (23) | Beta (α = 383.00; β = 305.00) | Dal Negro et al. (23) |

| Model input parameter | Base-case value | Source of base-case value range | DSA range | Source of DSA range | PSA parameters and distribution | Source of distribution parameter(s) |
|---|---|---|---|---|---|---|
| outpatient treatment, all age and risk groups | | | | | | |
| Mortality due to RSV-LRTD requiring hospitalization, including in-hospital and 1-month post-discharge mortality, by age group | | | | | | |
| 60-64 years | 8.0% | Estimated using the data reported by Puggina et al. (5) and adjusted to account for all-cause mortality (6) | 6.4% – 9.5% | ±20% of base-case value | Beta (α = 88.86; β = 1027.78) | Calibrated to the assumed +/-20% range** |
| 65-69 years | 17.8% | | 14.2% – 21.3% | | Beta (α = 78.79; β = 364.45) | |
| 70-74 years | 17.8% | | 14.2% – 21.3% | | Beta (α = 78.79; β = 364.45) | |
| 75-79 years | 34.6% | | 27.7% – 41.5% | | Beta (α = 62.44; β = 118.06) | |
| 80-84 years | 34.6% | | 27.7% – 41.5% | | Beta (α = 62.44; β = 118.06) | |
| 85+ years | 34.6% | | 27.7% – 41.5% | | Beta (α = 62.44; β = 118.06) | |
| Mortality due to RSV-LRTD requiring ED treatment, outpatient treatment, or no treatment, all age and risk groups | 0.0% | Assumption | Not varied | – | Not varied | – |
| Percentage of RSV-ARD cases with antibiotics use, by level of medical care, all age and risk groups | | | | | | |
| Hospitalization | 61.2% | Puggina et al. (5) | Not varied | – | Not varied | – |
| ED treatment | 77.0% | Santus et al. (24) | | | | |
| Outpatient treatment | 51.6% | Bracaloni et al. (25) | | | | |
| No treatment | 0.0% | Assumption | | | | |
| **All-cause mortality** | | | | | | |
| Death due to other causes (from 60 to 100 years) | Age-specific | General population mortality from | Not varied | – | Not varied | – |

| Model input parameter | Base-case value | Source of base-case value range | DSA range | Source of DSA range | PSA parameters and distribution | Source of distribution parameter(s) |
|---|---|---|---|---|---|---|
| | | 2025 Italian life tables (ISTAT) (6) | | | | |
| **Direct costs, all age and risk groups** | | | | | | |
| Acquisition cost of mRNA-1345 | €180 | Assumption in line with vaccine list price assumptions used in previously published Italian economic evaluations (26, 27) | Not varied | – | Not varied | – |
| Administration cost of mRNA-1345 | €7.47 | Capri et al. (15), ISTAT (16) | Not varied | – | Not varied | – |
| Cost per case of RSV requiring hospitalization | €4,282 | Estimated based on mean cost of DRGs 079, 080, 089, 090, 092, 093, 096, and 097 (28), ISTAT (16, 17) | €3,425.69 – €5,138.53 | ±20% of base-case value | Gamma (α = 96.79; θ = 44.24) | Calibrated to the assumed +/-20% range** |
| Cost per case of RSV requiring ED visit | €285 | Calabrò et al. (29), ISTAT (16) | €227.66 – €341.50 | ±20% of base-case value | Gamma (α = 96.79; θ = 2.94) | Calibrated to the assumed +/-20% range** |
| Cost per case of RSV requiring outpatient visit | €98 | Calabrò et al. (29), ISTAT (16) | €78.31 – €117.47 | ±20% of base-case value | Gamma (α = 96.79; θ = 1.01) | Calibrated to the assumed +/-20% range** |
| **HRQoL** | | | | | | |
| Baseline utility, by age group | | | | | | |
| 60-64 years | 0.907 | Scalone et al. (30) | 0.901 – 0.912 | CI estimated from Scalone et al. (30) | Beta (α = 8,357.83; β = 860.88) | Calibrated to the CI estimated from Scalone et al. (30) |
| 65-69 years | 0.892 | | 0.885 – 0.898 | | Beta (α = 7,738.91; β = 941.67) | |
| 70-74 years | 0.892 | | 0.885 – 0.898 | | Beta (α = 7,738.91; β = 941.67) | |

| Model input parameter | Base-case value | Source of base-case value range | DSA range | Source of DSA range | PSA parameters and distribution | Source of distribution parameter(s) |
|---|---|---|---|---|---|---|
| 75-79 years | 0.853 | | 0.844 – 0.863 | | Beta (α = 4,532.34; β = 779.83) | |
| 80-84 years | 0.853 | | 0.844 – 0.863 | | Beta (α = 4,532.34; β = 779.83) | |
| 85+ years | 0.853 | | 0.844 – 0.863 | | Beta (α = 4,532.34; β = 779.83) | |
| Life expectancy | Age- and sex-specific | ISTAT (6) | Not varied | – | Not varied | – |
| QALY loss per level of medical care, all age and risk groups | | | | | | |
| Hospitalization | 0.0193 | Hutton et al. (31, 32) | 0.0095 – 0.0316 | Range from Hutton et al. (31, 32) | Beta (α = 11.46; β = 582.46) | Calibrated to the CI reported by Hutton et al. (31, 32) |
| ED treatment | 0.0185 | Assumption | 0.0053 – 0.0347 | Assumption | Beta (α = 6.18; β = 327.71) | |
| Outpatient treatment | 0.0185 | Hutton et al. (31, 32) | 0.0053 – 0.0347 | Range from Hutton et al. (31, 32) | Beta (α = 6.18; β = 327.71) | |
| No treatment | 0.0093 | Assumption based on Moghadas et al. (33) | 0.0027 – 0.0174 | Assumption based on Moghadas et al. (33) | Beta (α = 6.25; β = 669.81) | |

* Individuals with at least one chronic condition were defined according to the PASSI d'Argento surveillance system as those with cardiovascular disease, stroke or cerebral ischemia, cancer (including leukemia and lymphoma), chronic respiratory diseases, diabetes, chronic liver disease/cirrhosis, or renal failure.

** Where PSA distribution parameters were not available from the literature used to inform the base-case, they were derived from the available evidence. Where only lower and upper bounds were available, distribution parameters were fitted by quantile matching such that the 0.025 and 0.975 quantiles corresponded to the lower and upper bounds, respectively.

*CI,* confidence interval, *DRGs,* diagnosis-related groups, *DSA,* deterministic sensitivity analysis, *ED,* emergency department, *HRQoL,* health-related quality of life, *PSA,* probabilistic sensitivity analyses, *QALY,* quality-adjusted life-year, *RSV,* respiratory syncytial virus, *RSV-ARD,* RSV-related acute respiratory disease, *RSV-LRTD,* RSV-related lower respiratory tract disease, *VE,* vaccine efficacy.

*Table S5. Scenario 1: High-Risk Adults Aged 60-74 Years. Detailed Results, Over 5 Years*

| Outcome | mRNA-1345 | No vaccination | Difference* |
|---|---|---|---|
| **Disease burden (number of cases, % cases avoided)** | | | |
| RSV-ARD cases (no LRTD) | 1,159,609 | 1,417,775 | -258,166 (-18%) |
| RSV-LRTD cases | 135,363 | 168,267 | -32,905 (-20%) |
| Antibiotic prescriptions | 406,179 | 498,084 | -91,905 (-19%) |
| Deaths | 4,296 | 5,229 | -933 (-18%) |
| **Healthcare resource use (number of cases, % cases avoided)** | | | |
| Hospitalizations | 20,025 | 24,714 | -4,689 (-19%) |
| ED visits | 4,778 | 5,939 | -1,161 (-20%) |
| Outpatient visits | 756,098 | 926,870 | -170,772 (-18%) |
| **Total QALYs lost** | **3,859,509** | **3,873,913** | **-14,404** |
| **Economic outcomes (€, millions)** | | | |
| Vaccination costs** | €537 | €0 | €537 |
| Hospitalization costs | €81 | €100 | - €19 |
| ED and outpatient costs | €72 | €88 | - €16 |
| **Total direct costs** | **€690** | **€189** | **€502** |
| **ICER, €/QALY gained** | | | **€34,816** |

* mRNA-1345 vaccine compared with no vaccine

** Including cost of vaccine acquisition, vaccine administration, and cost of adverse events

*ED,* emergency department, *ICER,* incremental cost-effectiveness ratio, *QALY,* quality-adjusted life-year, *RSV,* respiratory syncytial virus, *RSV-ARD,* RSV-related acute respiratory disease, *RSV-LRTD,* RSV-related lower respiratory tract disease.

*Table S6. Scenario 2: All Adults Aged ≥75 Years. Detailed Results, Over 5 Years*

| Outcome | mRNA-1345 | No vaccination | Difference* |
|---|---|---|---|
| **Disease burden (number of cases, % cases avoided)** | | | |
| RSV-ARD cases (no LRTD) | 1,165,326 | 1,430,671 | -265,345 (-19%) |
| RSV-LRTD cases | 304,022 | 385,262 | -81,240 (-21%) |
| Antibiotic prescriptions | 499,903 | 620,254 | -120,351 (-19%) |
| Deaths | 19,624 | 24,868 | -5,244 (-21%) |
| **Healthcare resource use (number of cases, % cases avoided)** | | | |
| Hospitalizations | 56,731 | 71,891 | -15,160 (-21%) |
| ED visits | 10,730 | 13,597 | -2,867 (-21%) |
| Outpatient visits | 885,281 | 1,096,208 | -210,927 (-19%) |
| **Total QALYs lost** | **12,767,953** | **12,801,026** | **-33,073** |
| **Economic outcomes (€, millions)** | | | |
| Vaccination costs** | €726 | €0 | €726 |
| Hospitalization costs | €232 | €295 | - €63 |
| ED and outpatient costs | €86 | €107 | - €21 |
| **Total direct costs** | **€1,044** | **€402** | **€642** |
| **ICER, €/QALY gained** | | | **€19,401** |

* mRNA-1345 vaccine compared with no vaccine

** Including cost of vaccine acquisition, vaccine administration, and cost of adverse events

*ED,* emergency department, *ICER,* incremental cost-effectiveness ratio, *QALY,* quality-adjusted life-year, *RSV,* respiratory syncytial virus, *RSV-ARD,* RSV-related acute respiratory disease, *RSV-LRTD,* RSV-related lower respiratory tract disease.

*Table S7. Scenario 3: Lower Bound For Vaccine Coverage. Detailed Results, Over 5 Years*

| Outcome | mRNA-1345 | No vaccination | Difference* |
|---|---|---|---|
| **Disease burden (number of cases, % cases avoided)** | | | |
| RSV-ARD cases (no LRTD) | 2,586,690 | 2,848,446 | -261,756 (-9%) |
| RSV-LRTD cases | 496,457 | 553,529 | -57,072 (-10%) |
| Antibiotic prescriptions | 1,012,210 | 1,118,338 | -106,128 (-10%) |
| Deaths | 27,008 | 30,097 | -3,089 (-10%) |
| **Healthcare resource use (number of cases, % cases avoided)** | | | |
| Hospitalizations | 86,681 | 96,605 | -9,924 (-10%) |
| ED visits | 17,522 | 19,536 | -2,014 (-10%) |
| Outpatient visits | 1,832,228 | 2,023,078 | -190,849 (-9%) |
| **Total QALYs lost** | **16,651,201** | **16,674,939** | **-23,738** |
| **Economic outcomes (€, millions)** | | | |
| Vaccination costs** | €631 | €0 | €631 |
| Hospitalization costs | €355 | €396 | - €41 |
| ED and outpatient costs | €176 | €195 | - €19 |
| **Total direct costs** | **€1,162** | **€591** | **€572** |
| **ICER, €/QALY gained** | | | **€24,078** |

* mRNA-1345 vaccine compared with no vaccine

** Including cost of vaccine acquisition, vaccine administration, and cost of adverse events

*ED,* emergency department, *ICER,* incremental cost-effectiveness ratio, *QALY,* quality-adjusted life-year, *RSV,* respiratory syncytial virus, *RSV-ARD,* RSV-related acute respiratory disease, *RSV-LRTD,* RSV-related lower respiratory tract disease.

*Table S8. Scenario 4: Higher Bound For Vaccine Coverage. Detailed Results, Over 5 Years*

| Outcome | mRNA-1345 | No vaccination | Difference* |
|---|---|---|---|
| **Disease burden (number of cases, % cases avoided)** | | | |
| RSV-ARD cases (no LRTD) | 2,063,179 | 2,848,446 | -785,267 (-28%) |
| RSV-LRTD cases | 382,312 | 553,529 | -171,217 (-31%) |
| Antibiotic prescriptions | 799,954 | 1,118,338 | -318,384 (-29%) |
| Deaths | 20,831 | 30,097 | -9,266 (-31%) |
| **Healthcare resource use (number of cases, % cases avoided)** | | | |
| Hospitalizations | 66,832 | 96,605 | -29,773 (-31%) |
| ED visits | 13,493 | 19,536 | -6,043 (-31%) |
| Outpatient visits | 1,450,529 | 2,023,078 | -572,548 (-28%) |
| **Total QALYs lost** | **16,603,724** | **16,674,939** | **-71,215** |
| **Economic outcomes (€, millions)** | | | |
| Vaccination costs** | €1,894 | €0 | €1,894 |
| Hospitalization costs | €272 | €396 | - €124 |
| ED and outpatient costs | €139 | €195 | - €56 |
| **Total direct costs** | **€2,305** | **€591** | **€1,715** |
| **ICER, €/QALY gained** | | | **€24,078** |

* mRNA-1345 vaccine compared with no vaccine

** Including cost of vaccine acquisition, vaccine administration, and cost of adverse events

*ED,* emergency department, *ICER,* incremental cost-effectiveness ratio, *QALY,* quality-adjusted life-year, *RSV,* respiratory syncytial virus, *RSV-ARD,* RSV-related acute respiratory disease, *RSV-LRTD,* RSV-related lower respiratory tract disease.

*Table S9. Scenario 5: Lower Bound Of 95% CI For Hospitalization Rate. Detailed Results, Over 5 Years*

| Outcome | mRNA-1345 | No vaccination | Difference* |
|---|---|---|---|
| **Disease burden (number of cases, % cases avoided)** | | | |
| RSV-ARD cases (no LRTD) | 2,428,237 | 2,978,663 | -550,426 (-19%) |
| RSV-LRTD cases | 336,500 | 423,926 | -87,426 (-21%) |
| Antibiotic prescriptions | 880,057 | 1,085,582 | -205,525 (-19%) |
| Deaths | 18,670 | 23,497 | -4,826 (-21%) |
| **Healthcare resource use (number of cases, % cases avoided)** | | | |
| Hospitalizations | 59,208 | 74,529 | -15,321 (-21%) |
| ED visits | 11,876 | 14,962 | -3,086 (-21%) |
| Outpatient visits | 1,617,181 | 1,992,614 | -375,433 (-19%) |
| **Total QALYs lost** | **16,596,905** | **16,636,870** | **-39,965** |
| **Economic outcomes (€, millions)** | | | |
| Vaccination costs** | €1,263 | €0 | €1,263 |
| Hospitalization costs | €242 | €305 | - €64 |
| ED and outpatient costs | €154 | €191 | - €36 |
| **Total direct costs** | **€1,659** | **€496** | **€1,163** |
| **ICER, €/QALY gained** | | | **€29,095** |

* mRNA-1345 vaccine compared with no vaccine

** Including cost of vaccine acquisition, vaccine administration, and cost of adverse events

*CI,* confidence interval, *ED,* emergency department, *ICER,* incremental cost-effectiveness ratio, *QALY,* quality-adjusted life-year, *RSV,* respiratory syncytial virus, *RSV-ARD,* RSV-related acute respiratory disease, *RSV-LRTD,* RSV-related lower respiratory tract disease.

*Table S10. Scenario 6: Higher Bound Of 95% CI For Hospitalization Rate. Detailed Results, Over 5 Years*

| Outcome | mRNA-1345 | No vaccination | Difference* |
|---|---|---|---|
| **Disease burden (number of cases, % cases avoided)** | | | |
| RSV-ARD cases (no LRTD) | 2,219,715 | 2,715,838 | -496,124 (-18%) |
| RSV-LRTD cases | 544,179 | 685,512 | -141,333 (-21%) |
| Antibiotic prescriptions | 932,589 | 1,151,694 | -219,105 (-19%) |
| Deaths | 29,238 | 36,784 | -7,545 (-21%) |
| **Healthcare resource use (number of cases, % cases avoided)** | | | |
| Hospitalizations | 94,612 | 119,064 | -24,451 (-21%) |
| ED visits | 19,206 | 24,195 | -4,988 (-21%) |
| Outpatient visits | 1,666,045 | 2,054,123 | -388,079 (-19%) |
| **Total QALYs lost** | **16,658,855** | **16,714,053** | **-55,198** |
| **Economic outcomes (€, millions)** | | | |
| Vaccination costs** | €1,263 | €0 | €1,263 |
| Hospitalization costs | €386 | €488 | - €101 |
| ED and outpatient costs | €161 | €199 | - €38 |
| **Total direct costs** | **€1,810** | **€687** | **€1,123** |
| **ICER, €/QALY gained** | | | **€20,348** |

* mRNA-1345 vaccine compared with no vaccine

** Including cost of vaccine acquisition, vaccine administration, and cost of adverse events

*CI,* confidence interval, *ED,* emergency department, *ICER,* incremental cost-effectiveness ratio, *QALY,* quality-adjusted life-year, *RSV,* respiratory syncytial virus, *RSV-ARD,* RSV-related acute respiratory disease, *RSV-LRTD,* RSV-related lower respiratory tract disease.

*Table S11. Scenario 7: Higher Bound For Cost Of RSV Management. Detailed Results, Over 5 Years*

| Outcome | mRNA-1345 | No vaccination | Difference* |
|---|---|---|---|
| **Disease burden (number of cases, % cases avoided)** | | | |
| RSV-ARD cases (no LRTD) | 2,324,935 | 2,848,446 | -523,512 (-18%) |
| RSV-LRTD cases | 439,385 | 553,529 | -114,144 (-21%) |
| Antibiotic prescriptions | 906,082 | 1,118,338 | -212,256 (-19%) |
| Deaths | 23,920 | 30,097 | -6,177(-21%) |
| **Healthcare resource use (number of cases, % cases avoided)** | | | |
| Hospitalizations | 76,757 | 96,605 | -19,849 (-21%) |
| ED visits | 15,508 | 19,536 | -4,029 (-21%) |
| Outpatient visits | 1,641,379 | 2,023,078 | -381,699 (-19%) |
| **Total QALYs lost** | **16,627,462** | **16,674,939** | **-47,477** |
| **Economic outcomes (€, millions)** | | | |
| Vaccination costs** | €1,263 | €0 | €1,263 |
| Hospitalization costs | €428 | €540 | - €112 |
| ED and outpatient costs | €555 | €687 | - €131 |
| **Total direct costs** | **€2,246** | **€1,227** | **€1,019** |
| **ICER, €/QALY gained** | | | **€21,465** |

* mRNA-1345 vaccine compared with no vaccine

** Including cost of vaccine acquisition, vaccine administration, and cost of adverse events

*ED,* emergency department, *ICER,* incremental cost-effectiveness ratio, *QALY,* quality-adjusted life-year, *RSV,* respiratory syncytial virus, *RSV-ARD,* RSV-related acute respiratory disease, *RSV-LRTD,* RSV-related lower respiratory tract disease.

*Table S12. Scenario 8: Alternative QALY Loss For Non-Hospitalized Cases. Detailed Results, Over 5 Years*

| Outcome | mRNA-1345 | No vaccination | Difference* |
|---|---|---|---|
| **Disease burden (number of cases, % cases avoided)** | | | |
| RSV-ARD cases (no LRTD) | 2,324,935 | 2,848,446 | -523,512 (-18%) |
| RSV-LRTD cases | 439,385 | 553,529 | -114,144 (-21%) |
| Antibiotic prescriptions | 906,082 | 1,118,338 | -212,256 (-19%) |
| Deaths | 23,920 | 30,097 | -6,177(-21%) |
| **Healthcare resource use (number of cases, % cases avoided)** | | | |
| Hospitalizations | 76,757 | 96,605 | -19,849 (-21%) |
| ED visits | 15,508 | 19,536 | -4,029 (-21%) |
| Outpatient visits | 1,641,379 | 2,023,078 | -381,699 (-19%) |
| **Total QALYs lost** | **16,619,000** | **16,664,492** | **-45,492** |
| **Economic outcomes (€, millions)** | | | |
| Vaccination costs** | €1,263 | €0 | €1,263 |
| Hospitalization costs | €313 | €396 | - €82 |
| ED and outpatient costs | €158 | €195 | - €37 |
| **Total direct costs** | **€1,734** | **€591** | **€1,143** |
| **ICER, €/QALY gained** | | | **€25,128** |

* mRNA-1345 vaccine compared with no vaccine

** Including cost of vaccine acquisition, vaccine administration, and cost of adverse events

*ED,* emergency department, *ICER,* incremental cost-effectiveness ratio, *QALY,* quality-adjusted life-year, *RSV,* respiratory syncytial virus, *RSV-ARD,* RSV-related acute respiratory disease, *RSV-LRTD,* RSV-related lower respiratory tract disease.

*Table S13. Scenario 9: Additional QALY Lost For Post-Hospitalization Period. Detailed Results, Over 5 Years*

| Outcome | mRNA-1345 | No vaccination | Difference* |
|---|---|---|---|
| **Disease burden (number of cases, % cases avoided)** | | | |
| RSV-ARD cases (no LRTD) | 2,324,935 | 2,848,446 | -523,512 (-18%) |
| RSV-LRTD cases | 439,385 | 553,529 | -114,144 (-21%) |
| Antibiotic prescriptions | 906,082 | 1,118,338 | -212,256 (-19%) |
| Deaths | 23,920 | 30,097 | -6,177(-21%) |
| **Healthcare resource use (number of cases, % cases avoided)** | | | |
| Hospitalizations | 76,757 | 96,605 | -19,849 (-21%) |
| ED visits | 15,508 | 19,536 | -4,029 (-21%) |
| Outpatient visits | 1,641,379 | 2,023,078 | -381,699 (-19%) |
| **Total QALYs lost** | **16,627,918** | **16,675,515** | **-47,597** |
| **Economic outcomes (€, millions)** | | | |
| Vaccination costs** | €1,263 | €0 | €1,263 |
| Hospitalization costs | €313 | €396 | - €82 |
| ED and outpatient costs | €158 | €195 | - €37 |
| **Total direct costs** | **€1,734** | **€591** | **€1,143** |
| **ICER, €/QALY gained** | | | **€24,017** |

* mRNA-1345 vaccine compared with no vaccine

** Including cost of vaccine acquisition, vaccine administration, and cost of adverse events

*ED,* emergency department, *ICER,* incremental cost-effectiveness ratio, *QALY,* quality-adjusted life-year, *RSV,* respiratory syncytial virus, *RSV-ARD,* RSV-related acute respiratory disease, *RSV-LRTD,* RSV-related lower respiratory tract disease.

*Table S14. Scenario 10: Shorter Time Horizon (3 years). Detailed Results, Over 5 Years*

| Outcome | mRNA-1345 | No vaccination | Difference* |
|---|---|---|---|
| **Disease burden (number of cases, % cases avoided)** | | | |
| RSV-ARD cases (no LRTD) | 1,408,645 | 1,804,478 | -395,834 (-22%) |
| RSV-LRTD cases | 260,523 | 348,020 | -87,496 (-25%) |
| Antibiotic prescriptions | 545,865 | 706,993 | -161,128 (-23%) |
| Deaths | 14,044 | 18,752 | -4,708 (-25%) |
| **Healthcare resource use (number of cases, % cases avoided)** | | | |
| Hospitalizations | 45,172 | 60,318 | -15,146 (-25%) |
| ED visits | 9,195 | 12,283 | -3,088 (-25%) |
| Outpatient visits | 990,330 | 1,279,947 | -289,617 (-23%) |
| **Total QALYs lost** | **10,420,988** | **10,459,092** | **-38,104** |
| **Economic outcomes (€, millions)** | | | |
| Vaccination costs** | €1,263 | €0 | €1,263 |
| Hospitalization costs | €190 | €253 | - €64 |
| ED and outpatient costs | €98 | €126 | - €29 |
| **Total direct costs** | **€1,550** | **€380** | **€1,170** |
| **ICER, €/QALY gained** | | | **€30,708** |

* mRNA-1345 vaccine compared with no vaccine

** Including cost of vaccine acquisition, vaccine administration, and cost of adverse events

*ED,* emergency department, *ICER,* incremental cost-effectiveness ratio, *QALY,* quality-adjusted life-year, *RSV,* respiratory syncytial virus, *RSV-ARD,* RSV-related acute respiratory disease, *RSV-LRTD,* RSV-related lower respiratory tract disease.

*Table S15. Scenario 11: Societal Perspective. Detailed Results, Over 5 Years*

| Outcome | mRNA-1345 | No vaccination | Difference* |
|---|---|---|---|
| **Disease burden (number of cases, % cases avoided)** | | | |
| RSV-ARD cases (no LRTD) | 2,324,935 | 2,848,446 | -523,512 (-18%) |
| RSV-LRTD cases | 439,385 | 553,529 | -114,144 (-21%) |
| Antibiotic prescriptions | 906,082 | 1,118,338 | -212,256 (-19%) |
| Deaths | 23,920 | 30,097 | -6,177(-21%) |
| **Healthcare resource use (number of cases, % cases avoided)** | | | |
| Hospitalizations | 76,757 | 96,605 | -19,849 (-21%) |
| ED visits | 15,508 | 19,536 | -4,029 (-21%) |
| Outpatient visits | 1,641,379 | 2,023,078 | -381,699 (-19%) |
| **Total QALYs lost** | **16,627,462** | **16,674,939** | **-47,477** |
| **Economic outcomes (€, millions)** | | | |
| Vaccination costs** | €1,263 | €0 | €1,263 |
| Hospitalization costs | €313 | €396 | - €82 |
| ED and outpatient costs | €158 | €195 | - €37 |
| Indirect costs | €686 | €859 | - €172 |
| **Total costs** | **€2,420** | **€1,449** | **€971** |
| **ICER, €/QALY gained** | | | **€20,447** |

* mRNA-1345 vaccine compared with no vaccine

** Including cost of vaccine acquisition, vaccine administration, and cost of adverse events

*ED,* emergency department, *ICER,* incremental cost-effectiveness ratio, *QALY,* quality-adjusted life-year, *RSV,* respiratory syncytial virus, *RSV-ARD,* RSV-related acute respiratory disease, *RSV-LRTD,* RSV-related lower respiratory tract disease.
